# Ultra-fast deep-learned pediatric CNS tumor classification during surgery

**DOI:** 10.1101/2023.01.25.23284813

**Authors:** C. Vermeulen, M. Pagès-Gallego, L. Kester, M.E.G. Kranendonk, P. Wesseling, J. van der Lugt, K. van Baarsen, E.W. Hoving, B.B.J. Tops, J. de Ridder

## Abstract

The primary treatment of CNS tumors starts with a neurosurgical resection in order to obtain tumor tissue for diagnosis and to reduce tumor load and mass effect. The neurosurgeon has to decide between radical resection versus a more conservative strategy to prevent surgical morbidity. The prognostic impact of a radical resection varies between tumor types. However due to a lack of pre-operative tissue-based diagnostics, limited knowledge of the precise tumor type is available at the time of surgery. Current standard practice includes preoperative imaging and intraoperative histological analysis, but these are not always conclusive. After surgery, histopathological and molecular tests are performed to diagnose the precise tumor type. The results may indicate that an additional surgery is needed or that the initial surgery could have been less radical. Using rapid Nanopore sequencing, a sparse methylation profile can be directly obtained during surgery, making it ideally suited to enable intraoperative diagnostics. We developed a state-of-the-art neural-network approach called Sturgeon, to deliver trained models that are lightweight and universally applicable across patients and sequencing depths. We demonstrate our method to be accurate and fast enough to provide a correct diagnosis with as little as 20 to 40 minutes of sequencing data in 45 out of 49 pediatric samples, and inconclusive results in the other four. In four intraoperative cases we achieved a turnaround time of 60-90 minutes from sample biopsy to result; well in time to impact surgical decision making. We conclude that machine-learned diagnosis based on intraoperative sequencing can assist neurosurgical decision making, allowing neurological comorbidity to be avoided or preventing additional surgeries.

## 1 Introduction

Central Nervous System (CNS) tumors are the most lethal category of tumors among children. Most commonly, the first line treatment of pediatric CNS tumors is neurosurgical resection of the tumor. During this procedure a delicate balance must be struck between maximizing resection on the one hand and minimizing the risk of neurological damage on the other hand (Duffau and Mandonnet 2013; Yong and Lonser 2011). An important factor in determining if the risk of a more aggressive resection is acceptable, is the tumor subtype (Cohen 2022). For instance, Diffuse Midline Gliomas with a Histone 3 mutation are considered incurable, indicating that surgery should primarily be aimed at acquisition of tumor tissue for diagnosis and preserving quality of life rather than attempting total resection (Karremann *et al*. 2018). Likewise, Medulloblastoma cases show limited prognostic improvement between total and near-total resection, also indicating that conservative resection is warranted (Thompson *et al*. 2016). In other cases radical resection is beneficial: in Posterior Fossa Ependymoma type A (PFE-A) a strategy of aiming at a Gross Total Resection (GTR) should be followed since this is an important prognostic factor (Venkatramani *et al*. 2012; Ramaswamy *et al*. 2016; Pajtler *et al*. 2017). In Atypical Teratoid Rhabdoid Tumor (ATRT) cases a similar trend was found where total resection improved overall patient survival (Egiz, Kannan, and Asl 2022). The neurosurgical strategy thus depends on a precise and reliable diagnosis of the tumor.

Current practice consists of preoperative imaging and intraoperative diagnosis achieved by rapid histological assessment of frozen tumor sections by a pathologist. However, these tests do not always result in a clear diagnosis, and are sometimes even revised based on postoperative tissue-based diagnostics. As a result, some patients require a second surgery, while others could in hindsight have been operated less radically. For this reason our aim is to have a fast and reliable method to classify CNS tumors during surgery and thereby to optimize surgical strategy for resection.

Altered genome-wide DNA methylation patterns are highly distinctive features of neoplasms, and the assessment of DNA methylation can reveal information about the origin and prognosis of a tumor (Papanicolau-Sengos and Aldape 2022). The most routinely used assays for diagnostic methylation profiling are the Illumina Infinium 450K and EPIC arrays (Capper, Jones, *et al*. 2018); Jaunmuktane *et al*. 2019; Priesterbach-Ackley et al. 2020), which interrogate cytosine methylation status of 450.000 (Sandoval *et al*. 2011) and 850.000 (Moran, Arribas, and Esteller 2016) CpG sites, respectively. Recent work demonstrated that methylation profiling can be used to accurately diagnose CNS tumors (Capper, Jones, *et al*. 2018; Jaunmuktane *et al*. 2019). Using machine learning approaches, in particular random forest classification, highdimensional methylation profiles can be accurately assigned to a specific CNS subtype (Capper, Jones, *et al*. 2018; Jaunmuktane *et al*. 2019). These methylation arrays in combination with the algorithm described by Capper *et al*. is widely used in routine diagnostic practice. However, the turnaround time for array-based methylation profiles is in the order of several days, even if the diagnostic workflow is optimized, and therefore incompatible with an intraoperative setting.

Nanopore DNA sequencing recently emerged as a method that enables ultrarapid sequencing-based diagnosis (Gorzynski *et al*. 2022; Sagniez *et al*. 2022). A major advantage of nanopore sequencing is that the sequencing data is available for analysis in real time. In addition, nanopore sequencing directly samples the native DNA strand thereby allowing direct measurement of methylated cytosines, significantly reducing sample preparation times (Xu and Seki 2020). Combined, these features make it ideally suitable for intraoperative methylation-based tumor classification. In this setting, a tissue sample is sent for sequencing in the early stages of surgery to obtain a molecular diagnosis in time to, combined with the histological diagnosis, affect and shape the neurosurgical strategy (Djirackor *et al*. 2021). A major challenge of this application is that in such a short time, only very sparse methylation profiles can be generated and the majority of potentially methylated sites is not covered by sequence reads. Moreover it is a priori unknown which sites will be covered.

To enable tumor classification using sparse and therefore rapidly obtainable data we have developed Sturgeon, a deep learning neural network classifier that is patient agnostic, is optimally tuned to deal with sparse data, and does not require in situ training or validation. In the Sturgeon approach, extensive computational resources can be allocated to train and validate highly performant and complex neural networks prior to surgery. This is a major advantage over existing classification algorithms that rely on patient-specific model training during surgery (Djirackor *et al*. 2021). Our final models are trained on 14 million and validated on 4 million simulated nanopore runs, respectively, which is practically impossible to achieve with in situ training due to the computational demands. After training and validation, the resulting Sturgeon model is portable and only takes a few seconds to run on a laptop CPU. This allows us to extensively validate the robustness of our models on independent datasets, from which we simulate tens of thousands of sequencing runs. Our approach thus allows training and applying multiple models, fine tuning for different classification tasks, and to apply them in parallel in a clinically relevant timeframe.

As a proof of concept we trained Sturgeon models for CNS tumor classification, and retrospectively applied them on sparse nanopore sequencing data in 48 pediatric CNS tumor samples and 415 publicly available nanopore sequenced CNS samples. The model shows very high accuracy and is able to correctly classify the vast majority of patients (45 out of 49) with the equivalent of 20-40 minutes of sequencing, in line with a 90 minute time-window between biopsy and diagnosis. Finally, we demonstrate the ability of Sturgeon to influence surgical decision making by applying it in a realistic intraoperative setting for four independent pediatric CNS tumor resection surgeries.

## 2 Results

### 2.1 Data augmentation and simulation enables effective neural network training

To reach a turn around time (TAT) of 60-90 minutes only very limited nanopore sequencing data can be generated, in the order of 100-400 Mb. As a result, extremely sparse coverage across the entire genome is expected (covering 0.5-4% of the CpG sites in a 450K array), and it is a priori unknown which sites will be covered. This poses a significant challenge for the downstream machine learning model. As large well-annotated nanopore-based methylation datasets are currently lacking and will take years to reach the comprehensiveness of the available array-based datasets, we developed a simulation strategy that generates realistic training data from array-based methylation profiles (or other methylation measurement data). Finally, effectively training neural network models requires orders of magnitude more training samples than the number of patient samples available. Sturgeon therefore employs a data augmentation approach to effectively upsample the number of training samples available. This approach also allows for class-balancing by upsampling small classes relatively more compared to larger classes.

Sturgeon is designed to train a neural network on simulated nanopore sequencing runs from the publicly available Infinium 450K profiles reported in Capper *et al*. (Capper, Jones, *et al*. 2018). This dataset contains 2801 reference labeled methylation profiles from CNS tumor samples. The simulation consists of the following components (**Figure 1**): (1) Binarization of the array beta values, to account for the fact that in the sparse setting, where the maximum coverage is 1x, heterogeneously methylated sites cannot be detected. (2) Non-uniform CpG site sampling to account for the fact that nanopore sequence reads are 5Kb in the rapid sample prepping methods used in an intraoperative setting. (3) Variable sampling of the number of CpG sites covered, to account for read accumulation as time progresses. (4) Random error, to account for the fact that nanopore methylation calling has an expected error rate of 10% according to the most performant methylation caller (Megalodon combined with the Rerio CpG methylation model, as suggested in (Yuen *et al*. 2021)).

**Fig. 1.**
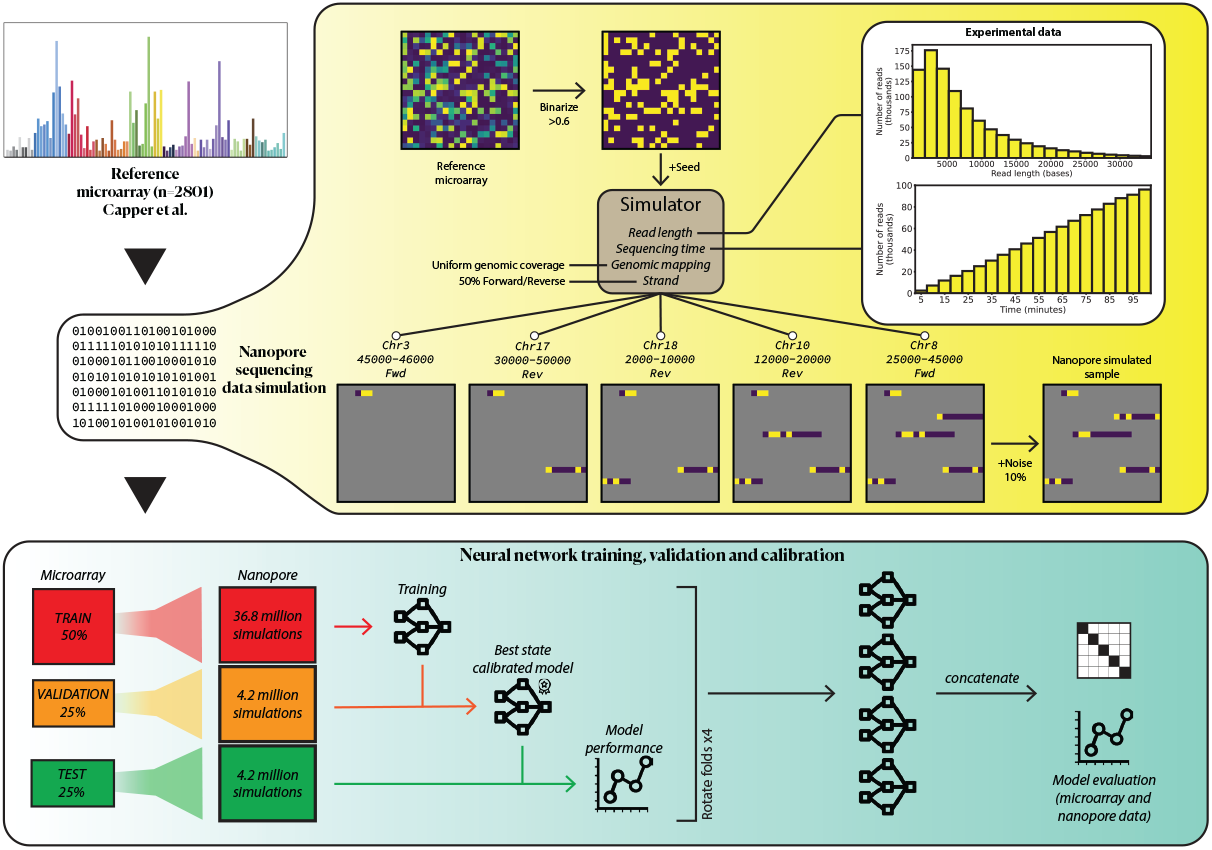
Schematic representation of the simulation and cross-validation processes. Capper *et al*. reference dataset. with 2801 CNS samples is used for training, validation (assessing best performing state and calibration) and testing (final model performance). We simulate nanopore sequencing runs based on previous existing sequencing runs (read length distribution and throughput), since these simulations produce very sparse samples we can simulate millions of samples. We perform 4-fold cross-validation, and rotate the folds to obtain 4 models that are used in the final prediction of external microarray data and nanopore sequencing data.

The resulting model consists of four neural networks (submodels), each trained, validated and calibrated independently (**Figure 1**). To this end, we split the Capper *et al*. reference dataset into 4 folds while keeping the original class distributions. We then use two folds to train the submodel, one fold to determine the best performing state of the submodel and to perform score calibration and the final fold to evaluate the submodel’s performance. Simulations are tightly controlled through the seeds of the pseudo-random number generator, i.e. training, validation and test seeds are mutually exclusive, to avoid cross-validation leakage. We rotated folds between submodels in order to independently incorporate and evaluate the whole reference dataset (**Figure 2**). New samples are then classified by all four submodels, and the result of the most confident submodel is reported.

**Fig. 2.**
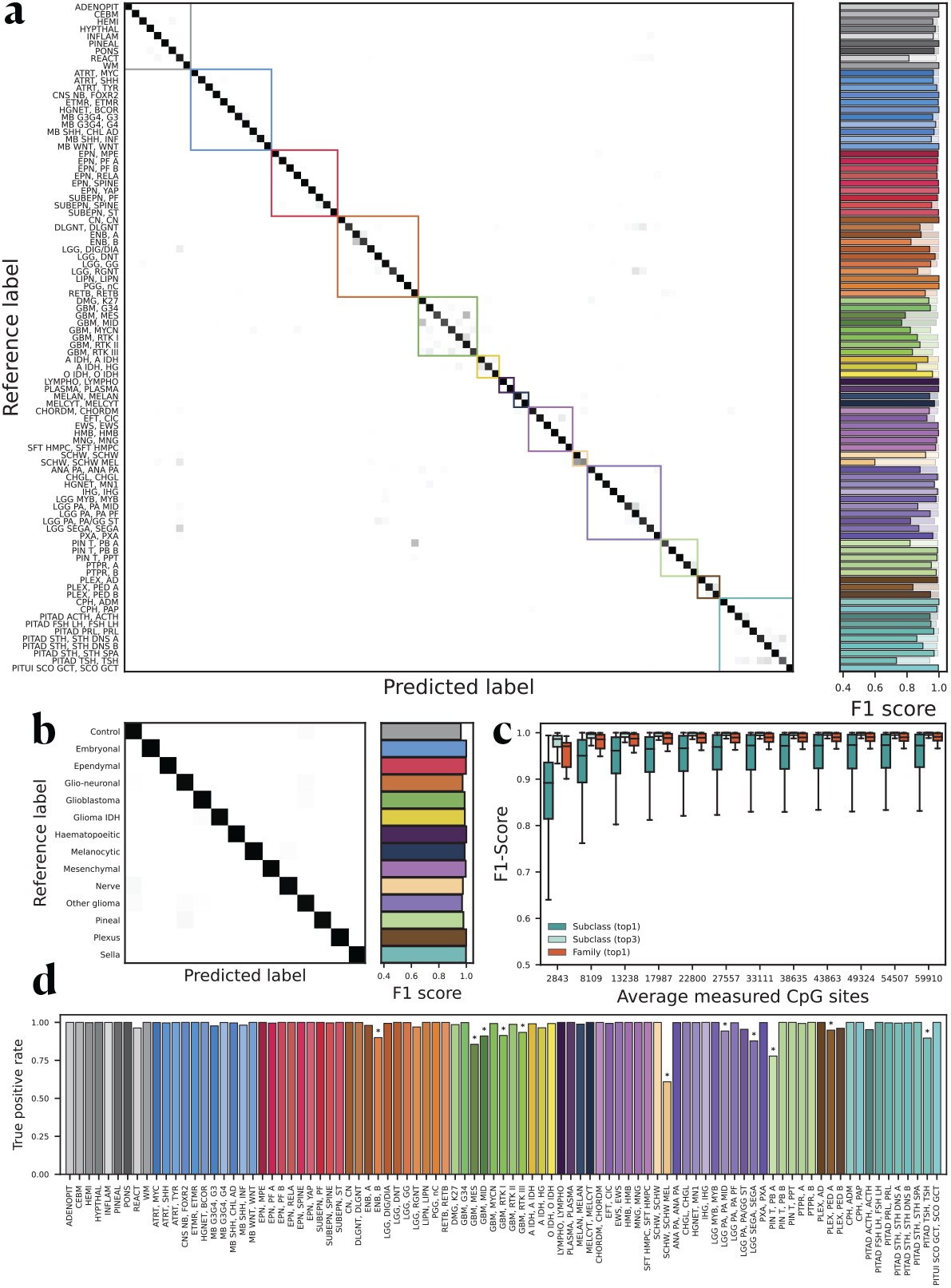
Sturgeon submodel cross-validated evaluation. Sturgeon performance on the four test folds of the Capper *et al*. dataset. (a) Confusion matrix showing the highest scoring class for each reference label at 40 minutes of simulated sequencing (~97% missing values from microarray data). Bars on the right side of the plot indicate the top 1 (solid) and top 3 (transparent) F1-scores per reference label class. (b) Performance when scores are aggregated per family label. (c) F1-scores at increasing sequence depths per class and per family, and when taking into account the top 3 classifications. (d) True positive rate for each class at the 0.95 score threshold.

### 2.2 Sturgeon submodels achieve *>*0.94 true positive rate within 40 minutes of simulated sequencing

We first evaluated Sturgeon submodels based on left-out array-based methylation data. From each sample in the test fold we simulated 500 sparse samples at each of twelve different simulated sequencing depths (from 0.6% to 14% coverage of the reference data; **Supplementary Figure 1-2 and Supplementary Table 2**). Applying Sturgeon to these data shows that at approximately 40 minutes of simulated sequencing, the submodels reach an average F1-score of 0.935 across all classes (**Figure 2a**). Some confusion remains between Melanotic Schwannoma (0.589 F1-score) and regular Schwannoma samples; between TSH-secreting pituitary adenomas (0.716 F1-score) and other pituitary adenomas; and also some confusion is observed between glioblastoma subclasses (0.847 average F1-score). When considering the top 3 classifications, an average 0.992 F1-score across all classes (ie. in the majority of cases the correct label is in the top 3 of highest classifications) is reached; and performance on these aforementioned difficult classes becomes comparable to the rest (**Figure 2a**). Notably, the few misclassifications are predominantly confusions of samples within the same family; when we aggregate scores for subclasses within families, the average F1-score for family classification is 0.984 (**Figure 2b**). As expected, Sturgeon’s performance is directly correlated to the sequencing depth and confidence improves as more sequencing data is available. However, most significant improvements occur within the (simulated) first 50 minutes of sequencing, with the number of covered CpG ranging from 0.6% to 4% of the 450K available sites (**Figure 2c, Supplementary Figures 1-2**).

We next sought to calibrate the classifiers, meaning that a classification score of 0.7 should translate to a 70% chance of a correct classification. For this purpose we applied temperature scaling (Guo *et al*. 06–11 Aug 2017). After calibration the overall Expected Calibration Error (ECE), decreased from 0.023 to 0.003 in the validation set and from 0.025 to 0.002 in the test set (**Supplementary Table 3, Supplementary Figures 3-6**). As this does not solve the challenges of sparse calls at intermediate confidence scores, we decided to conservatively use a cut-off score of 0.95 to confidently classify a sample. Using this cut-off, 80 out of the 91 classes in the test set have a True Positive Rate (TPR) higher than 0.95; for a less conservative threshold of 0.8, 26 classes do not reach the expected TPR (**Figure 2d, Supplementary Figure 7**). The 11 classes that do not reach the 0.95 threshold have a TPR between 0.8 and 0.9, except for Melanotic Schwannoma. However, the presence of melanin pigment in melanotic schwannoma (an exceedingly rare tumor, renamed in the WHO 2021 CNS tumor classification as malignant melanotic nerve sheath tumor) can be expected to allow for discriminating these tumors from ordinary schwannomas based on histological evaluation.

### 2.3 Sturgeon performance compared to the Heidelberg V11b4 classifier in pediatric samples

The training dataset for Sturgeon consists of a varied patient population of different ages, which is not necessarily a good representation of the expected population in a pediatric oncology center. We therefore aimed to validate Sturgeon on pediatric methylation profiles. For this purpose we obtained 94 EPIC profiles generated for patients that underwent a brain tumor resection surgery in the Princess Maxima Center for Pediatric Oncology. For each of these samples, the publicly available “Heidelberg classifier (V11b4)” was applied in routine clinical care and results were used in clinical decision making. This classifier can be considered an updated version of the Capper *et al*. classifier since it is based on an extremely comprehensive collection of EPIC profiles. For the analysis of Sturgeon performance we divided the samples based on the output of the Heidelberg classifier. The “clear diagnosis” group is defined as samples where the Heidelberg classifier reached a score of 0.84 or higher (N=68); the cutoff recommended by the developers of the Heidelberg classifier (Capper, Stichel, *et al*. 2018). The “difficult diagnosis” group is defined as samples with a score below the 0.84 cutoff (N=26). This group is considered difficult to diagnose based on methylation profile, which is likely to occur for uncommon tumor types that do not correspond to any of the previously annotated classes, tumors that occur in the context of a genetic tumor syndrome, heterogeneous samples or samples with a low tumor purity.

For each methylation profile, we simulate 500 nanopore sequencing runs at seven sequencing depths, as described before, for a total of 332.500 simulated nanopore sequencing experiments, after which we applied the Sturgeon classifier (**Figure 3, Supplementary Table 1, supplementary figures 8-9**).

**Fig. 3.**
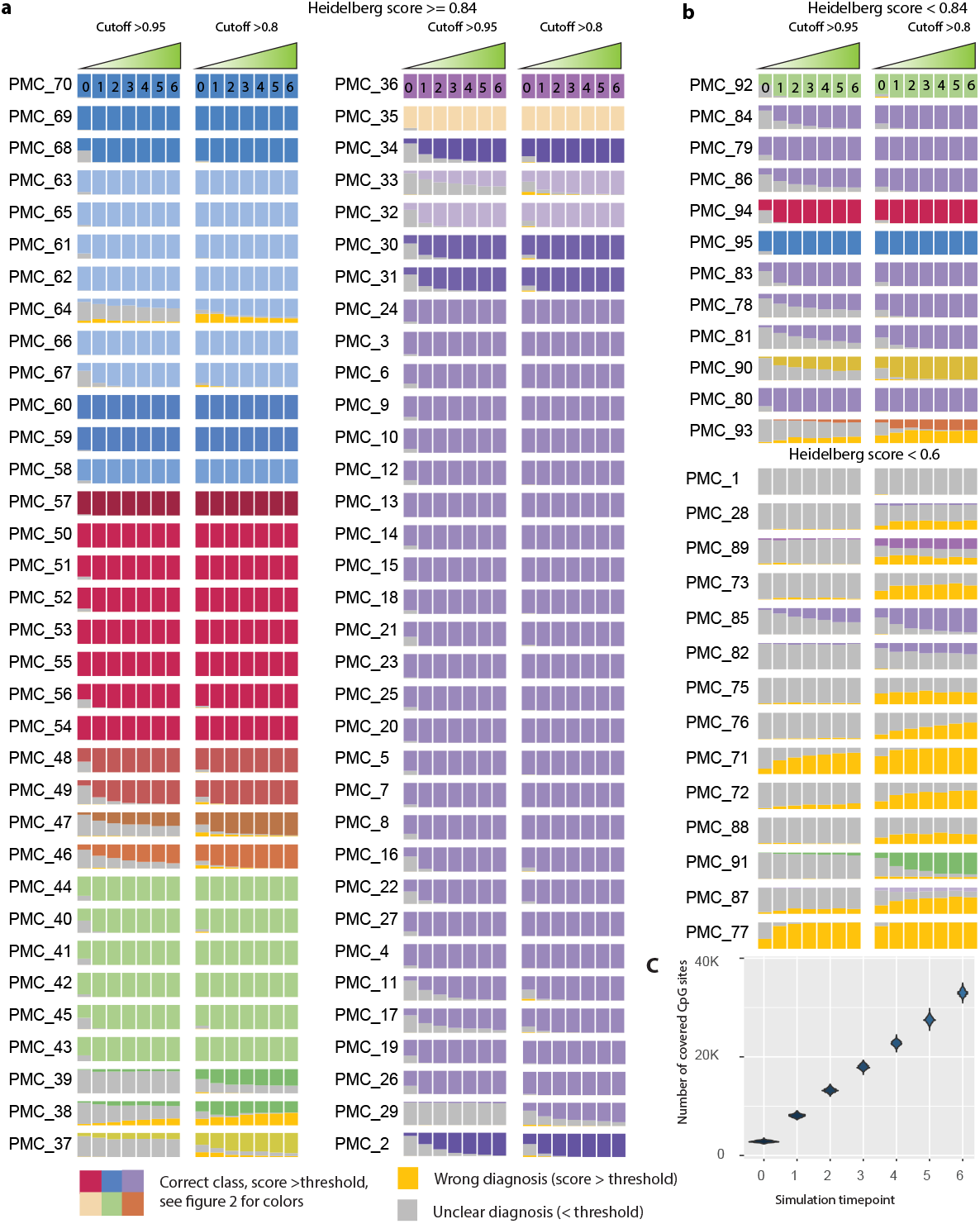
Classification performance over time on nanopore runs simulated from pediatric CNS tumor methylation arrays. For each of 96 methylation profiles, a series of nanopore sequencing experiments were simulated. At each timepoint 500 experiments were simulated corresponding to approximately 5 minutes of sequencing per timepoint. Each bar indicates a consecutive timepoint and simulated sequencing data is accumulated over time. A stacked bar graph is plotted based on the number of correct, unclear or wrong classifications. Correct classifications are those with a confidence score *>*0.95 (left) and *>*0.8 (right) and with a class corresponding to the true diagnosis (bars are colored according to the class label). Unclear classifications are those with confidence-scores *<*0.95 or *<*0.8 colored in gray). Wrong classifications are misdiagnoses where a confidence-score *>*0.95 or *>*0.8 is obtained for the incorrect class (colored in yellow). (a) Clear diagnosis group (Heidelberg classifier score *>*0.84) (b) Difficult diagnosis group (Heidelberg classifier score *<*0.84). (c) Distribution of the number of CpG sites covered at each simulated timepoint.

We consider the classification result at two thresholds. A score exceeding 0.95 indicates confident diagnosis that can be provided to the pathologist. A score exceeding 0.8 indicates a likely diagnosis which can also be communicated but with some caution, if time permits further sequencing is recommended as the confidence will likely increase. We consider scores below 0.8 unsafe to communicate with the clinical staff, and more data needs to be obtained. For the clear diagnosis group Sturgeon classified correctly (at the 0.8 threshold) in 95.3%(32412 of the 34000 simulated samples) in as little as 25 minutes of simulated sequencing (timepoint 1, **figure 3c**, average of 8091 (1.7%) 450K CpG sites covered). For the conservative threshold of 0.95, still 86.2% (29316/34000) of simulated samples were correctly classified. At the same time point, only 2.7% and 13.8%of simulations did not reach a confidence score exceeding 0.8 and 0.95 respectively. Wrong diagnoses were called in 2.0% of simulations at the 0.8 threshold, and only 0.5% for the conservative 0.95 threshold.

At 50 minutes of simulated sequencing (timepoint 3 in **Figure 3c**, an average of 17945 CpG sites covered), performance improved slightly, with 97.1% (33020/34000) simulations reaching a correct diagnosis with confidence *>*0.8 and 90.8% with a score over 0.95. 1.6% did not reach a score *>*0.8. Wrong diagnoses were called in only 1.3% of simulations with a score over 0.8 and 0.5% with a score *>*0.95. Taken together, these results suggest that a conclusive diagnosis can be reached within 25-50 minutes of sequencing for the vast majority of pediatric cases that can be classified using the heidelberg v11b4 classifier, with a very low false positive rate.

For the cases where the Heidelberg classifier was not able to provide a diagnosis (N=26), Sturgeon was also less performant in general. For most of these cases a definitive diagnosis was reached based on the combination of molecular and histological features. In 11 of the 27 cases Sturgeon frequently reached a diagnosis in concordance with the pathologist’s diagnosis (but often at later time points). All of these cases also reached a Heidelberg classifier score between 0.6 and 0.84 (**see Figure 3b and Supplementary Table 1**). In the other cases, both sturgeon and the Heidelberg classifier performed poorly, most frequently resulting in an unclear diagnosis (low confidence scores or high scores for control tissue classes). This can be attributed to different reasons; Four samples (PMC_1, PMC_28, PMC_82 and PMC_76) had a Low tumor fraction based on histology.

Four samples were diagnosed with a class not present in the 2018 classification scheme. PMC_71; low grade glioma with PLAG-FOXO fusion (Sievers *et al*. 2021), sometimes classified by sturgeon as a SUBEPN. PMC_88; low grade glioma with a PLAG amplification (Keck *et al*. 2022), Sturgeon does not reach the 0.8 threshold in this sample, but we note that the highest scoring class is often Medulloblastoma Group 3. PMC_77; High grade glioma with underlying Li-Fraumeni syndrome consistently classified as a Glioblastoma with a MYCN amplification across different simulations. Similarities between MYCN amplified and Li-Fraumeni tumors have recently been reported (Guerrini-Rousseau *et al*. 2023). We theorize that the methylation profile could be similar between a TP53 loss and a MYCN amplification, as MYCN directly opposes TP53 function (Agarwal *et al*. 2018). PMC_73: a low grade biphenotypic glioneuronal tumor sometimes classified as a Pilocytic Astrocytoma/Ganglioglioma).

Two samples could not be classified by histology either and to date do not have a definitive diagnosis (PMC_72 and PMC_75). Finally four of the underperforming cases originate due to germline mutations (PMC_89, PMC_85, PMC_91 and PMC_77 (Li-fraumeni mentioned before)) which has previously also been suggested to complicate methylation-based classification (Jaunmuktane *et al*. 2019).

Together these results indicate that Sturgeon can perform on par with the Heidelberg v11b4 classifier, even when only a very limited number of (simulated) nanopore sequence reads is available. It also reiterates the limitation that Sturgeon (as any other machine learning-based classifier) is only able to perform well in samples that are sufficiently represented in the training data. Reassuringly, for classes that are not represented in the training data, confidence scores are usually low, resulting in an unclear outcome rather than a misdiagnosis.

### 2.4 Sturgeon provides accurate diagnoses from sparsely nanopore sequenced samples

To assess the performance of Sturgeon in a realistic setting, we retrospectively sequenced and classified 26 pediatric brain tumor DNA samples obtained from the Princess Maxima center biobank. We then applied Sturgeon to increasing numbers of reads, simulating a normal minION sequencing run in 5 pseudotime (see methods) minute intervals (**Figure 4b**).

**Fig. 4.**
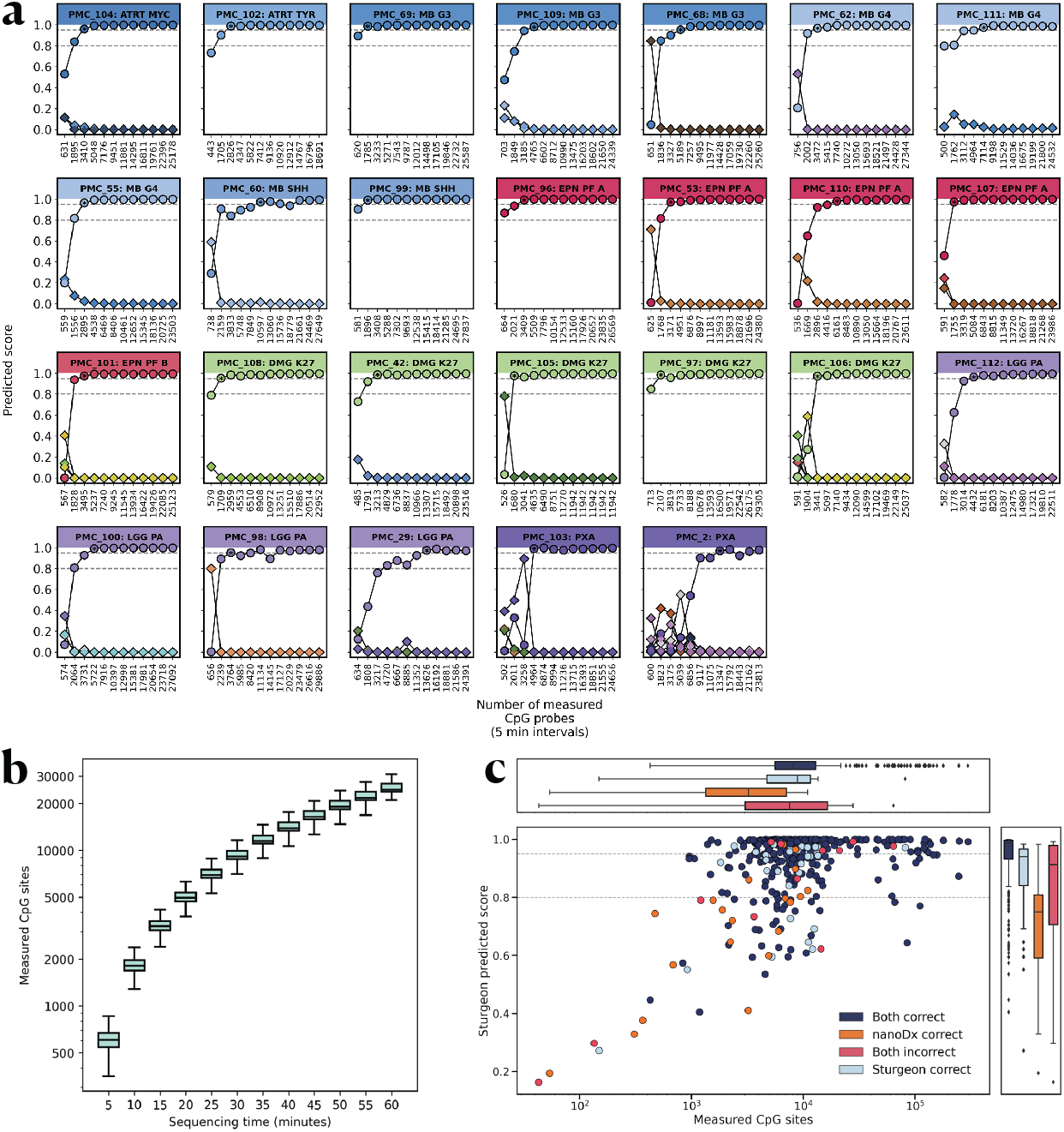
Sturgeon applied to nanopore sequenced samples. (a) Sturgeon classification scores for 26 pediatric CNS tumor samples at increasing sequencing time (5 minute pseudo time intervals). Top bar indicates the sample name and diagnosis. Circles indicate the predicted score of the correct class; diamonds indicate the predicted score of incorrect classes (classes with overtime averaged scores lower than 0.1 are omitted). Asterisks indicate the first time point where the score of the correct class was higher than 0.8. Horizontal dashed lines indicate the 0.8 and 0.95 thresholds. (b) Distributions for the number of covered CpG sites from the reference microarray data at increasing sequencing durations in nanopore sequencing MinION runs. (c) Sturgeon classifications on 415 CNS nanopore sequenced samples from a publicly available dataset (GSE209865), compared to the results from the nanoDx classifier. Either both classifiers were correct (dark blue), only nanoDx was correct (orange), only Sturgeon was correct (light blue), or both classifiers were incorrect (red). Horizontal dashed lines indicate the 0.8 and 0.95 score thresholds.

The classification results demonstrate that for 23 out of 26 samples Sturgeon assigned a score higher than 0.95 to the correct class after the equivalent of 25 minutes of sequencing; and on average such threshold was achieved between 15-20 minutes of sequencing (**Figure 4a, Supplementary table 4**). Samples PMC_60, PMC_29 and PMC_2 reached the 0.95 threshold at 30, 40 and 40 minutes of sequencing respectively. Samples PMC_68, PMC_98 and PMC_103 had an incorrect score higher than 0.8, but were correctly predicted in the following time points and the correct class score stabilized afterwards.

For each sample we generated 200.000 reads (3.9Gb), where a typical intraoperative run (60 minutes of sequencing) would be expected to yield 60.000 reads (200Mb) of throughput. This allowed us to evaluate the robustness of the results by randomly subsampling sequence reads, essentially simulating a different order in which the DNA molecules were sequenced. Our results show that Sturgeon is very robust, reporting the correct class in 29252 (23984 with score *>*0.95, 27354 with score *>* 0.8) out of the 31200 predictions (each sample is simulated 100 times and predicted 12 times (5 pseudotime minute intervals) per simulation). Notably it only reports the incorrect class 11 and 263 times with scores higher than 0.95 and 0.8 respectively (**Supplementary Table 5**). With increasing data available, the outcomes are more confident and accurate (**Supplementary figure 11**). It also showcases how some samples are easier to classify than others. For example, samples PMC_2 and PMC_29, which took longer than average to classify (**Figure 4a**), still have simulations in which the 0.95 threshold is not reached.

We next validated Sturgeon on a publicly available dataset (GSE209865) generated by Kuschel *et al*. (Kuschel *et al*. 2021), consisting of nanopore sequencing data for 415 CNS tumor samples available (**Figure 4c, Supplementary Table 6**). We note that the number of sequencing reads per sample is highly variable for this dataset and that for 24 samples fewer than 2000 CpG sites are covered, a number expected with less than 15 minutes of sequencing in a MinION flowcell (**Figure 4b**). We find that Sturgeon outperforms nanoDx, the patient-specific random forest classifier, as it is able to correctly predict 9 additional samples (**Supplementary figure 12**). Sturgeon correctly classified 383 (92.2%) samples, 343 (82.6%) at a threshold of 0.8 and 252 (60.7%) samples with a confidence *>*0.95. From the 415 samples, 32 (7.7%) were incorrectly classified of which 13 (3.1%) reached a confidence *>*0.8 and 8 (1.9%) reached a confidence score *>*0.95. Interestingly, on samples with fewer than 2000 CpG sites covered, Sturgeon still managed to correctly classify 14 samples, 3 of which at the 0.95 confidence threshold; however, the remaining 10 the samples were classified incorrectly. Overall, Sturgeon performs better and is more confident with an increasing number of measured CpG sites; and nanoDx is able to perform better on low CpG scenarios due to its patient-tailored model.

### 2.5 Site-specific classification can further improve turnaround time

The Capper *et al*. dataset consists of 80 classes, however, many class distinctions are only relevant within a particular topological context. Thus prior to surgery many classes can be ruled out. For instance, for a surgery of the spinal mass, a classifier does not need to be able to detect pituitary adenomas. We reasoned that by merging such irrelevant classes from the training dataset into a single class (“Other Non Brainstem”), the model can focus on the truly relevant classes and improve its performance. Compared to other regions in the brain, the number of relevant classes in the brainstem is relatively low (N=21,**Supplementary Table 8**); and to test this hypothesis, we developed a Sturgeon classifier specifically for brainstem tumors. We trained the model on the complete Capper *et al*. dataset, however, non-brainstem classes were grouped together as a single class (“Other Non Brainstem”) with the exception of the control classes.

We first evaluate the performance of the brainstem classifier on the pediatric EPIC profiles through cross-validation, as described earlier. Expectedly, for samples that are diagnosed with a class that is included as a brainstem class in the classifier training and with a clear heidelberg classifier score (*>*0.84, N=56), the brainstem classifier reaches high confidence scores at early timepoints (Supplementary Table 7). When using a conservative threshold confidence of 0.95, at timepoint 1 25461 out of 28000 (91.0%) of the simulations reach a definitive diagnosis, and 1953 (7.0%) of simulations are unclear (ie the classifier reports a score *<*0.95 or a high score for control tissue or classified as “Other”). At timepoint 3, the number of definitive diagnoses decreases slightly to 25260 (90.0%) and the number of unclear diagnoses is increased to 2131 (7.6%). The number of wrong diagnoses is 586 (0.6%) and 609 (0.7%) at timepoints 1 and 3 respectively. This indicates that the brainstem classifier reaches an optimal performance at very early timepoints compared to the general classifier. We note that practically all of the misdiagnoses arise from two of the 35 samples: PMC_38 and PMC_63. PMC_38 is a Glioblastoma GBM G34 that seems difficult to classify and is frequently mis-classified as a Glioblastoma GBM MID, similar classifications happen in the general classifier (**see figure 3**). PMC_63 is a Glioblastoma - GBM - Group 4 that is frequently misclassified as a Glioblastoma - GBM - Group 3 (again in both the general and brainstem classifiers).

In samples that did not reach the 0.84 threshold in the heidelberg classifier, but that were diagnosed as a class within the brainstem classifier (N=15), results were mixed: 4199 out of 7500 (55.0%) simulations were correct at timepoint 1, and 3274 (43.7%) were unclear. The brainstem classifier only made 27 misclassification out of 7500 simulations at timepoint 1 (**Supplementary Table 7**). When applying the brainstem classifier to samples from classes that do not typically occur in the brainstem (N=23), Sturgeon results in unclear outcomes (other or low scores) in the vast majority of cases, except in PMC_77 where it consistently classifies the Li-Fraumeni tumor as a Glioblastoma with MYCN amplification as discussed before.

To further evaluate the performance of the brainstem classifier, we obtained 24 additional tumor DNA samples from the PMC_biobank that originated in the brainstem (two of which; PMC_42 and PMC_105 were also included in the previous analyses). Reflective of the epidemiology of pediatric brainstem tumors, the vast majority of samples (23/24) were diagnosed with a glioblastoma harboring a H3K27 alteration. We multiplex sequenced these samples in two batches on the PromethION platform to a depth of *>*100.000 reads per sample. We then evaluated the performance of both the brainstem-specific and general classifier on an increasing number of covered CpG sites, with a rate expected in a single-sample MinION run (**Supplementary Figure 5, Supplementary Figure 11, Methods**). The general classifier slightly underperformed compared to earlier samples, as expected from the initial training and validation where glioblastoma subclasses showed a reduced TPR (**Figure 1A, Figure 1d**), it did not reach a confident diagnosis (score *>*0.95) in 4/24 samples and misclassified one sample (PMC_116) with a 0.84 confidence score. In comparison, the brainstem-specific classifier resulted in higher confidence score at earlier time points in most but not all cases (**Figure 5a, Supplementary Figure 14**), but also resulted in an unclear classification (“Other non brainstem” location classification) in 5/24 samples. The brainstem classifier correctly classified PMC_116. Notably the brainstem classifier seems prone to changing its classification to the “Other non brainstem” class at later time points (for example PMC_44, PMC_121, PMC_126), likely due to the high diversity within that particular training class. Notably in three out of five cases where the EPIC array classification was unclear (score below 0.84), a diagnosis was reached by the brainstem classifier (PMC_122), or both the brainstem and general classifier (PMC_121, PMC_114). In cases where both models reached the 0.95 threshold, the brainstem classifier was faster in 15 cases (saving on average 10 minutes of sequencing); however, it delayed diagnosis in one case by 20 minutes. We finally tested the brainstem classifier on non-brainstem samples. The classifier performed as good as the general classifier, and properly classified non-occurring brainstem classes as “Other location” (**Supplementary Figure 15**).

**Fig. 5.**
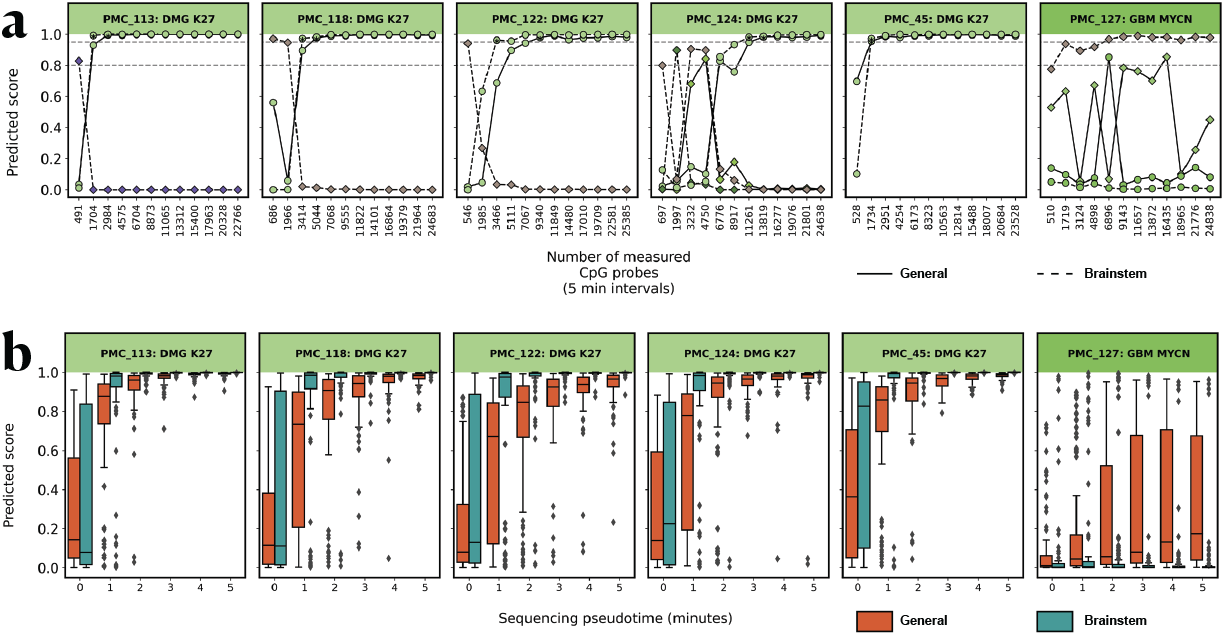
Results of the brainstem classifier compared to the general classifier. (a) Reads were accumulated in the order they were obtained at a rate based on the average minION sequencing speed. At each (approximately 5 minutes) timepoint both the brainstem and general classifier were applied, only correct classification classes are displayed (brainstem: dashed line, general: full line). Asterisks indicate the first time point the predicted score is higher than 0.95. Horizontal dashed lines indicate the 0.8 and 0.95 score thresholds. Mixed color labels indicate the diagnosis class that would be correct for the general (left) and brainstem (right) classifiers. (b) Reads were randomly selected from all sequence reads obtained per sample at a rate based on an average minION sequencing run. For each timepoint 100 random samplings were performed and the brainstem and general classifier were applied (brainstem: green, General: orange). Boxplots indicate median and interquartile range.

We further assessed the added value of the brainstem classifier by generating 100 random read samplings per time point and comparing the scores between the brainstem and the general classifier (**Figure 5b, Supplementary Figure 16**) Overall, the brainstem classifier achieves higher scores during early sequencing; for example, on average, the correct class scores are 0.20, 0.33 and 0.22 points higher during the first 5, 10 and 15 minutes of sequencing respectively. Furthermore, the brainstem classifier reaches the 0.95 score on average in ∼ 15 minutes, while the general classifier in∼ 20 minutes. A similar trend can also be observed on samples that are not from the brainstem, but that fall within the brainstem classes (**Supplementary Figure 17**).

To mitigate the risk of a misdiagnosis due to overfitting, or when an unusual tumor type is encountered in the brainstem, and to take advantage of both classifiers, the brainstem classifier can easily be deployed in parallel to the general Sturgeon classifier. In which case a high brainstem score should always be supported by a similar class from the general classifier, but possibly with a less stringent confidence threshold in the general classifier.

### 2.6 Intraoperative sequencing is compatible with surgical timeline

To demonstrate the clinical feasibility of Sturgeon in an intraoperative sequencing context, we performed the protocol intraoperatively on 4 samples. Samples obtained for histological assessment during surgery were split, and one part was used for intraoperative sequencing while the other part was used for histological assessment. In our institute, roughly 15 minutes are required to move the sample from the operating room to tissue processing where an appropriately sized sample is cut from the surgical sample. We note that this time can be further reduced by positioning the laboratory closer to the operating room. We optimized our DNA extraction protocol to rapidly obtain a high concentration of input DNA. In brief: we heavily reduced the lysis duration, relying on Qiashredder columns instead, and shortened all centrifugation times (see methods). This enables us to isolate DNA from brain tumor samples within 17-20 minutes. The library preparation is completed in approximately 15 minutes after DNA isolation, and sequencing commences roughly 30 minutes after the sample is received in the DNA isolation lab.

The sequencing itself is slowed down by the startup phase (**Supplementary Figure 11**), where mostly sequencing adapters are sequenced, and then ramps up towards higher pore activity and more informative reads per minute. Nevertheless, we typically obtain 10.000-20.000 sequence reads within one hour after the samples arrive in the isolation lab. In some, but not all, cases this is enough for a reliable diagnosis. After an hour and 30 minutes we typically obtain 40.000-60.000 reads, which is enough to produce a reliable diagnosis for the majority of retrospective and simulated cases as described above.

To demonstrate feasibility we applied the method intraoperatively in four cases (PMC_130 PMC_134) (**Supplementary Figure 18** shows the results of a slightly older model used at the time of sequencing, and **Supplementary Figure 19** shows the result with the most recent model). Due to the validation status of this method we did not communicate any outcomes with the clinical staff. In all cases sequencing was initiated within 40 minutes of the sample arriving in the DNA isolation lab.

In the first case (PMC_130) the tumor fraction was too low for analysis. Based on histology, this sample was classified as an Adamantinomateuos Craniofaryngioma, and it was also not submitted for an EPIC array due to the low tumor purity. Sturgeon did not reach a certainty threshold for any of the tumor classes.

In the second case (PMC_131) the pre-operative imaging pointed to a high likelihood of a medulloblastoma or ependymoma. After 30 minutes of sequencing, Sturgeon classified the sample as a G3 Medulloblastoma (non-WNT, non-SHH) with a score *>*0.8. Histological assessment also pointed towards a likely medulloblastoma and this was communicated to the operating room. Following the surgery, an EPIC array was also generated and submitted to the heidelberg classifier, this confirmed the diagnosis of Medulloblastoma Group 3 (v11b4, score 0.99). In this case the Sturgeon outcome could have provided the additional confirmation needed in time to adjust the neurosurgical strategy, in this case to opt for a relatively conservative resection as this yields a similar prognosis to GTR (Thompson *et al*. 2016).

In the third case (PMC_132) the sample submitted for nanopore sequencing did not contain tumor cells but a blood clot. DNA isolation yield was unusually low, and sequencing was compromised. Sturgeon did not report a score over 0.8 at any time point.

In the fourth case (PMC_133) the diagnosis was already known (Schwannoma located in the spine), but was not communicated to the scientific personnel. The entire procedure for this case was timed and filmed (**Figure 6a, Supplementary Video, Supplementary Figure 13**). One hour and 22 minutes after receiving a call from the operating room, the sample was classified by Sturgeon as a Schwannoma with a score over 0.8. This diagnosis was also confirmed using an EPIC array (v11b4, score 0.99).

**Fig. 6.**
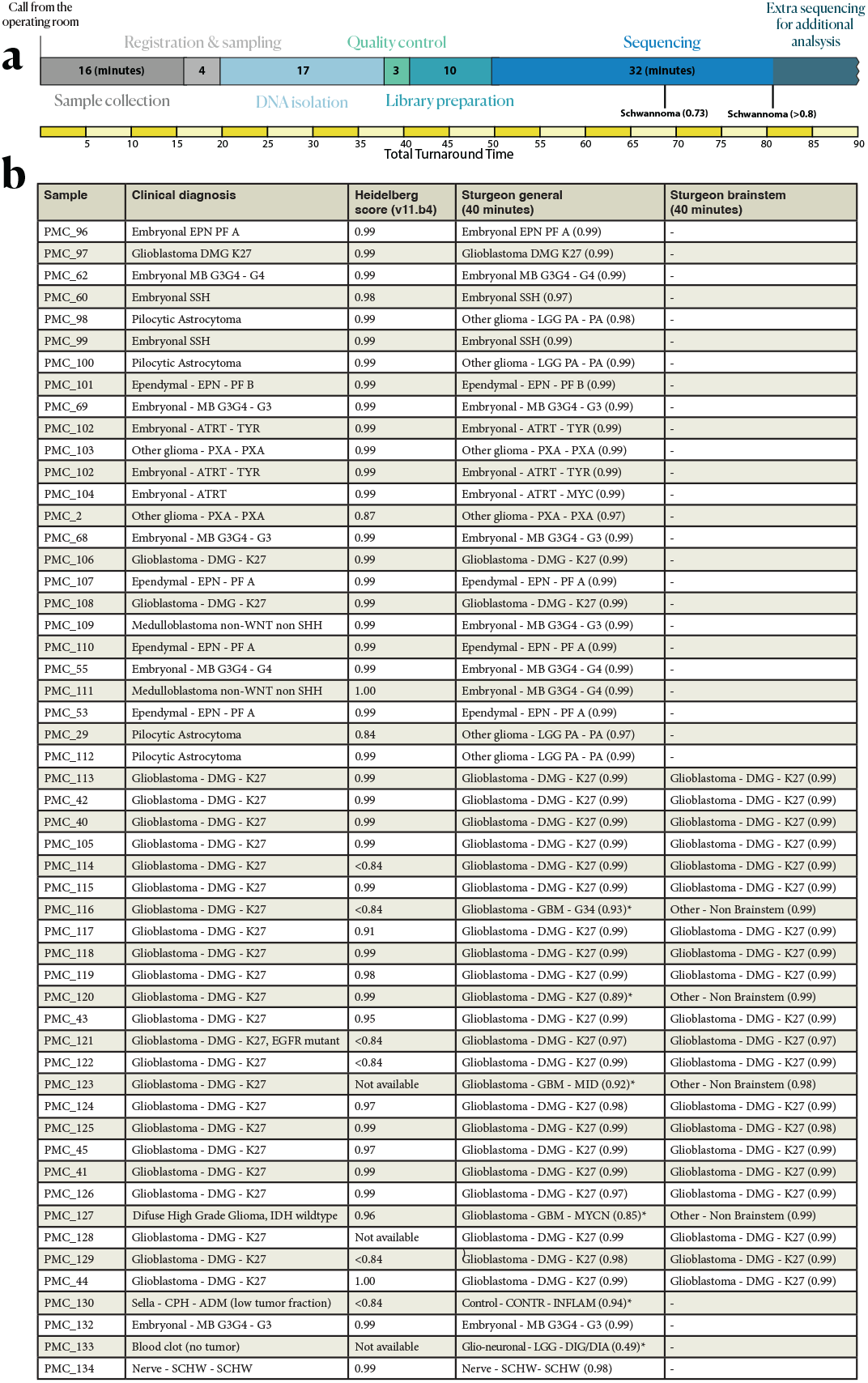
Intraoperative run timing and summary of all nanopore sequenced classified samples. (a) Timeline for the PMC_134 intraoperative run (as recorded in Supplementary Video) with the total turnaround time and required time per processing step indicated. (b) Table summarizing all patients where nanopore sequencing was performed. Clinical diagnosis refers to the diagnosis reached using best available means, including EPIC arrays, CNV profiling and histology. Heidelberg score indicates the score reached when the EPIC data (if generated) was classified by the heidelberg v11b4 classifier. Sturgeon outcome and Sturgeon Brainstem (if applicable) indicate the class and highest score obtained during 40 minutes of sequencing (* samples with a confidence score below 0.95).

Together these cases show that this approach can consistently yield clinically valuable information within 90 minutes from the moment a sample is obtained (**Figure 6b**), providing pathologists with an orthogonal diagnostic tool to assess the tumor class and ultimately, potentially preventing unnecessary surgical comorbidity or the need for a second surgery.

## 3 Discussion

Here we demonstrate the practical feasibility of intraoperative methylation-aware nanopore sequencing for pediatric brain tumor subtyping. Previously,the challenges of intraoperative sequencing have been addressed using a random forest approach, wherein the classifier is trained and validated during the surgery, based on the covered sites obtained at that point in time (Djirackor *et al*. 2021). Training a classifier “on the fly” requires a large and thus potentially time consuming computational effort during the surgery and also requires retraining at each timepoint. The resulting classifier is unique to the patient, and is therefore not reusable for other patients. Validation and score-calibration of the classifier needs to occur during the surgery, and may suffer from overfitting and therefore require on-site machine learning expertise. Finally, full methylation profiles of a large collection of patients have to be available on-site during training, meaning that every center that seeks to apply this method needs to have a copy of all the training data. Distributing such large patient datasets may not be compatible with privacy laws or patient consent.

To address these challenges, we developed Sturgeon, a deep learning approach that, despite being trained on methylation arrays, can still accurately classify tumor types based on the very sparse information obtained from time-constrained nanopore sequencing. Sturgeon uniquely moves the computationally intensive model training, validation and calibration phase outside the surgical time window, providing well-tested highly accurate one-size-fits-all classifications. Sturgeon models are not patient specific and can be used universally without retraining, mitigating the need to have access to privacy sensitive training data at the site of deployment. As a result, only limited computational resources are required during surgery. For example, the Sturgeon classifiers shown here can classify a megalodon output file containing data from 32610 reads in 17 seconds on a AMD Ryzen 7 6800H CPU. As the classification step practically poses no constraint on the time it takes to classify a sample, it is possible to run multiple Sturgeon classifiers in parallel. We show that the models perform robustly across different sequencing devices (minION and PromethION), laboratories (Utrecht and Berlin) and methylation calling methods (Megalodon and Nanopolish). However, as any other methylation classifier to date, performance is limited by tumor purity in the analyzed sample, and cannot account for intratumor heterogeneity.

We envision training of improved versions of Sturgeon as more data becomes available. The class definition used in the Capper *et al*. data, used for training Sturgeon, has since been updated several times, with the addition of many new classes (WHO Classification of Tumours Editorial Board 2022). However, to our knowledge, no up-to-date training dataset is available to the community for these new classes and therefore Sturgeon is unable to discern them. Furthermore the 450K arrays used by Capper *et al*. have since been surpassed by EPIC arrays with nearly twice as many CpG sites. Training Sturgeon on EPIC datasets would thus increase the information density per sequence read, and likely shorten the time to diagnosis. Furthermore, with nanopore-based methylation sequencing becoming a real alternative to array-based techniques, more sizable patient cohorts with nanopore-based methylation profiling may become available. Including such truly genome-wide datasets in the model training will make them more robust as more sites in the genome can be leveraged to make a prediction in the sparse situation.

Leveraging all available data for training Sturgeon is complicated due to data sharing restrictions as a result of privacy legislation that follows from the patient’s consent. Sturgeon is ideally suited to address this as it can readily be employed in a federated learning setting. Herein, rather than collecting training data at a central location, the model is distributed to participating institutes for training with local data for model refinement. Models are then returned to a central point to incorporate model updates after which the model is redistributed. Moreover, due to the simulation approach employed by Sturgeon, we envision that different types of training data, such as Infinium (450K or EPIC), nanopore or bisulfite sequencing data can all be naturally accommodated.

Ultra rapid methylation sequencing may hold great potential for several other fields of application. An increasing number of tumor types are routinely analyzed and/or diagnosed using EPIC arrays. Typical turnaround times for these arrays is four days or more, which can cause undesirable patient distress and anxiety. The Sturgeon approach may be a straightforward approach to drastically reducing the diagnostic TAT in these cases. Moreover, the required equipment cost is low; a consumer grade computer and a minION device, amounting to a combined investment of 2-3 thousand euros, can already be sufficient. This low capital investment also makes the application ideally suited for diagnostics in settings where finances are limited or where no dedicated pathologists are available. Return on investment may further be increased by considering that nanopore sequence reads can also be used to identify copy number variation (CNV) profiles. While this would require increased sequencing depth compared to the intraoperative application described here, the sequencing device can continue sequencing after a methylation-based diagnosis is reached. Sufficient coverage can easily be obtained using a single flowcell, thus providing very cost-effective whole-genome methylation profiling as well as CNV profiling for additional diagnostic consideration.

When implementing the Sturgeon approach in clinical practice, further improvements can be considered. For instance, sequencing can be performed on two flowcells simultaneously. This can further speed up sequencing time, by pooling the data from both flowcells, or aid in preventing wrong diagnoses due to “sampling errors”, by treating the data from both (or more) flowcells as duplicates. Alternatively, larger (but more expensive) sequencing devices such as PromethION, are available and could similarly improve the speed or reliability of the method.

In conclusion our results demonstrate that TATs of one and a half hours are feasible for the majority of samples. This is fully compatible with the timelines of conventional intraoperative histological assessment of the sample. We envision clinical application of Sturgeon to be deployed parallel to histological assessment by a trained pathologist who then also integrates the results into a final verdict. Using Sturgeon in this way could also reduce the requirement for a confidence score of *>*0.95 since the pathologist will always weigh the predicted tumor class in the context of the observed tumor histology. Sturgeon can thereby play an important role in guiding decision making in challenging cases where the histological picture is ambiguous. As a result, Sturgeon offers tangible potential to prevent second surgeries or surgical comorbidity and improve patients’ quality of life.

## 4 Methods

### 4.1 Data simulation

Short nanopore sequencing runs yield low (*<*1X) and random coverage of the genome. To enable model training, we generate simulated sparse nanopore runs based on the microarray data. To this end, N simulated reads are randomly sampled from the read length distribution (D) and assigned a start mapping position in the genome. N and D are defined based on a nanopore whole genome sequencing run (**Figure 1**). Forward or reverse direction is chosen at random. Reads are clipped at the start/end of the chromosome. Given this set of reads, the covered CpG sites are determined and their binarized methylation status is obtained from the microarray sample. To include measurement noise due to the nanopore methylation calling error rate, 10% of the covered CpG sites are randomly flipped. To reduce overtraining on specific sparsity levels, we simulate runs ranging from 5 to 60 minutes of sequencing in 5 minute intervals and combine the samples of different sparsity levels in a balanced fashion in a single training set (see below). To ensure reproducibility and avoid simulation leakage between samples of the different cross-validation folds, simulations can be completely deterministic (with the exception of noise) given a random seed and the simulation time.

### 4.2 Cross-validation

To assess model performance the Capper *et al*. dataset (Capper, Jones, et al. 2018) is split in four equally sized class-stratified folds. Two folds are used for submodel training, one for validation to assess the best model state during training and to perform score calibration. The final fold is used for testing to assess the submodel performance. Folds are rotated so that a total of four submodels are obtained. Simulation seeds are kept separate between the three folds: we used seed values between 0-499 for the test fold, between 500-999 for the validation fold and between 1000-1001000 for the training fold.

### 4.3 Submodel training

Sturgeon is a neural network containing three fully connected layers. The first two layers have 256 and 128 dimensions respectively, and are followed by a sigmoid activation. The last linear layer has a dimensionality equal to the number of classes to be predicted (91 dimensions for the general classifier, and 30 dimensions for the brainstem classifier). Dropout rate between layers was set to 0.5. As classification loss cross-entropy with uniform weights was chosen. We train the neural network in a two step process: we first pretrain a neural network on the Capper *et al*., 2018 classes (91 classes) using simulations that range between 0.6% and 14% sparsity. We then finetune this neural network for the final classifier by training using simulations that range between between 0.6% and 6.3% sparsity. For the brainstem classifier, the last layer is substituted by an untrained layer with the correct dimensionality (30 classes).

We pretrain the initial neural network for a total of 3000 epochs with a batch size of 256. For this purpose the AdamW optimizer (Loshchilov 2019) is used with a starting learning rate of 1e-5 that increases linearly for the first 1000 training batches until 1e-3; afterwards, it is decreased using a cosine function until it reached 1e-4 on the 1000th epoch; we then keep training at a constant learning rate of 1e-4 for 2000 epochs. Other parameters of the optimizer are: *β*_1_ = 0.9, *β*_2_ = 0.999, *ϵ* = 1e-8 and *λ* = 0.0005. We define one epoch as the number of reference samples in the most abundant class multiplied by the number of output classes. For every 2000 training batches the current weights of the model are saved; and the model is evaluated on 50 validation batches (12.800 samples) by calculating their average loss and sensitivity. Validation batches are sampled in the same manner as the training batches, with the exception that simulation seeds were independent. We finetune the neural network using the exact same parameters as described for the pretraining, with the exception that we finetune for 3000 epochs with a constant learning rate of 1e-4.

During inference, we classify samples using the four trained submodels and use as final classification the scores from the model with the highest confidence.

### 4.4 Adaptive Sample Balancing

Because of class imbalance in the training dataset, all classes are upsampled such that they are equally represented by simulating additional samples for classes smaller than the largest class. Similarly, we balance the sequencing sparsity levels such that the training data for each class consists of samples that have a uniform distribution of simulated sequencing times. At the end of each epoch, we recalculate the class balance by increasing the upsampling of classes and/or simulation times for which the model performs worse. Conversely, classes and/or simulation times for which the model performs well are upsampled relatively less. The number of samples for each class (c) and sparsity level (t) for epoch i+1 is provided by:

**Fig. 7.**
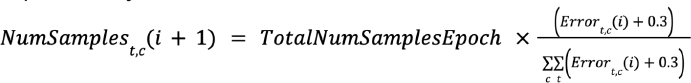
Adaptive sampling function.

We add a correction constant (0.3), to avoid completely removing classes or timepoints from the epoch. The total number of samples per epoch is kept constant, based on the first epoch.

### 4.5 Score calibration

To calibrate the classifier scores, enabling interpretation of these scores as confidence scores, we use temperature scaling (Guo *et al*. 2017). To optimize the calibration, we simulate each validation fold sample sample 500 times (using all validation fold seeds) for all sparsity levels (between 0.6% and 14% sparsity). Given the whole reference dataset, 2801 samples, this results in 16.806.000 total simulations. Based on these data we optimize the temperature parameter. To this end the non-scaled logits output from the last layer of the network are used to minimize the class weighted cross-entropy after dividing the non-scaled logits by the temperature parameter. For this purpose we use the L-BFGS algorithm implemented in PyTorch with learning rate 0.01 and a maximum of 500 iterations. We evaluate the calibration of the model using the Expected Calibration Error (ECE), a statistical measure that summarizes the difference between classifier accuracy and confidence. The ECE is defined as the weighted average of the absolute difference between accuracy and confidence on equally sized bins B (here we use 10 bins).

**Fig. 8.**
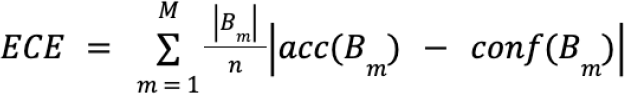
Score calibration function.

### 4.6 Model evaluation

We assessed the final performance of the model on the left-out test-fold samples. For this purpose, each sample was simulated 500 times and for all sparsity levels. In this way, each sample contributes 6000 simulated samples to the test set. We report top1 and top3 F1-scores for each class individually across all time intervals, as well as average metrics across classes.

### 4.7 Pseudotime

To reduce costs, some Nanopore sequencing samples used for validation were multiplexed in a MinION or PrometheION flowcell. Multiplexed sequencing times are not directly comparable to sequencing runs of a single sample. Similarly, samples sequenced on PrometheION flowcells are not directly comparable to MinION flowcells due to their larger throughput. In order to make these runs comparable to a real intraoperative scenario (one sample and one MinION flowcell), we use the number of Megalodon CpG calls as a proxy for sequencing time. This also avoids skewing the data for samples with shorter or longer than average read lengths. We use the median amount of CpG calls per 5 minute time bin from all MinION sequencing runs as the expected sequencing throughput (**Supplementary Figure 11**). The number of CpG sites collected from 5 to 60 minutes in increments of 5 minutes is: 51924, 104073, 124078, 149111, 173504, 194399, 207456, 217193, 232101, 241278, 247600, 258197). Thus, in a multiplexed sequencing run, we take the first N reads until the expected CpG calls are reached for that time bin. Furthermore, because the number of CpG calls is directly correlated with sequencing throughput (**Supplementary Figure 11**), differences in read length distribution are not affected by this. Finally, this also allows us to properly simulate the ramp up in sequencing throughput that happens during the first minutes of sequencing.

### 4.8 Robustness analysis

To analyze the robustness of the results on our Nanopore sequencing runs, we randomize the order of the sequenced reads and simulate 12 consecutive 5 minute (pseudotime) sequencing bins. We randomize the sequenced read order of each sample 100 times and evaluate in which time bin the desired threshold would have been reached and whether the classification was correct or not.

### 4.9 Methylation profile validation

EPIC arrays are routinely performed on pediatric CNS cancer samples. We gathered 95 such profiles that were generated in the routine diagnostic process within the princes Máxima pediatric cancer center. Raw EPIC profiles were binarized with a cutoff of beta *>*0.6 using scripts kindly provided by the authors of (Kuschel *et al*. 2021). EPIC probes were downsampled to match those within the 450K set (the number available in the reference cohort). We then simulate 500 nanopore runs at 12 sparsity levels as described above. The EPIC profiles were all submitted to the heidelberg v11b classifier (with the exception of PMC_20 which was classified with classifier v12.5), results (classification and score) thereof are listed in **Supplementary table 7**. Samples were also labeled with a “final diagnosis”, the result of a combination of histological assessment, imaging, CNV profiling and molecular characterization which we consider the ground truth.

### 4.10 Classification of publicly available nanopore sequencing data

We downloaded nanopore sequencing data from GSE209865. Of note, this dataset consists of processed sequencing data, which uses a different processing method, using Guppy and nanopolish, and mapping to hg19 (Kuschel *et al*. 2021), which can result in a reduced number of CpG calls compared to using megalodon combined with a rerio model (Yuen *et al*. 2021); but also results in binary methylation calls for EPIC methylation probes and can thus directly be used for Sturgeon classification.

### 4.11 DNA extraction and library prep

DNA is extracted from a tumor sample using an adapted Qiagen QiaAmp mini protocol. A tumor sample of roughly 5×5×5mm is (ideally) used as input material. 180uL ATL buffer is added to the sample and the sample is shortly ground using a pestle, then 200uL buffer AL and 20uL Proteinase K are added and the sample is moved to a 70 degree heat-block. Once heated the sample is ground with a pestle every minute to improve proteinase K accessibility. When the sample contains no more solid tissue or after five minutes of incubation/grinding, the sample is added to a Qiashredder column (Qiagen ID: 79656), not including any solid matter if still present. Shredder column is centrifuged at 20.000xG for 1 minute. 200uL of 96% ethanol is added to the eluate and the eluate is moved to a qiaAmp column and centrifuged for 1 minute at 6000xg. The column is washed with 500uL AW1, centrifuged at 6000g for 1 minute, then with 500uL AW2 at 12.000xg for 1.5 minute. Remaining ethanol is removed in a fresh elution column, centrifuged at 12.000xg for 30 seconds. Sample is eluted with 25 uL of MilliQ water. Samples are quantified using a nanodrop. Samples are library prepped using the Oxford Nanopore RBK004 kit using 600 ng input material and following manufacturers instructions for other steps.

### 4.12 Flowcell loading

ONT MinION sequencing initializes with a pore scan, which takes around 5 minutes and produces no sequence reads. Therefore we start the sequencing as soon as the sample arrives in the lab, so that sequencing commences as soon as the library is loaded onto the flowcell. Flowcells are primed using 800uL Flush Buffer (from ONT flowcell priming kit) at the start of the DNA isolation, after five minutes the flowcell is flushed with 200uL flush buffer and sequencing is initiated at which point the software will first perform a pore scan. The DNA library is loaded as soon as it is ready, at which point the pore scan has typically finished and actual sequencing commences.

### 4.13 Methylation calling

To call methylation from nanopore data we use Megalodon V2.5.0, which runs with Guppy V5 to perform basecalling and mapping to the CHM13 reference genome. To call per-read-per-site methylation we use the Rerio CpG methylation model as described by Yuen *et al*. (Yuen *et al*. 2021). Methylation calls are collated to the 450K CpG sites using 50 base pairs windows centered on the CpG site targeted by each infinium probe. If multiple CpG sites are present within the 50 base pairs window, majority voting is used to convert the calls to a single call per read. When multiple reads cover the same infinium probe site, majority voting is also used to create a methylation call. The methylation calling error rate was evaluated by comparing the methylation calls between nanopore sequencing and the microarray data for the same samples where both methods were available. This indicated a consistent concordance of 88-90% between binarized array data (beta cutoff at *>*0.6) and nanopore methylation calls (**Supplementary Table 1**). The error rate is evenly distributed between false positives (calling unmethylated sites as methylated) and false negatives (calling methylated sites unmethylated).

### 4.14 Live analysis

We developed a custom R script to run parallel to the sequencing software. As we noticed that megalodon outperforms the default guppy methylation calling, we run minKNOW with disabled basecalling. MinKNOW outputs fast5 files, each containing 4000 reads. The R script checks the minKNOW output folder for new fast5 files, and checks if they contain 4000 reads. Complete fast5 files are copied to a separate working directory where they are processed using megalodon. qCat is used to identify barcodes and depending on user settings either the most frequent or a user-specified barcode is selected. “Per-readper-site” methylation calls originating from reads with the selected barcode are saved and sturgeon is run. Sequencing and analysis are performed on an ASUS TUF A15 FA507RR-HN003W laptop with 64Gb RAM. All live analysis, including the experiment registered on film, were performed with an earlier version of the general model (V1), that had not been as extensively trained as the model presented in this work (V2). Results for the live analysis with the older model version can be found in **Supplementary Figure 18**, and results for the new model version can be found in **Supplementary Figure 19**.

## Data Availability

Data will be made available via the EGA repository upon publication.

## Acknowledgments

We acknowledge the Utrecht Sequencing Facility (USEQ) for providing sequencing service and data. USEQ is subsidized by the University Medical Center Utrecht and The Netherlands X-omics Initiative (NWO project 184.034.019).

## Declarations

- **Funding:** this project was partially funded by the Oncode Institute technology development fund.
- **Conflict of interest:** JdR, MPG and CV are inventors on a patent covering the development of Sturgeon. JdR is co-founder and director of Cyclomics, a genomics company.
- **Consent for publication:** The research was approved by the Biobank and Data access committee (BDAC) of the Princess Máxima Center for pediatric oncology. All included patients provided written informed consent for participation in the biobank (International Clinical Trials Registry Platform: NL7744). Inclusion of intraoperative samples was ethically approved via the same decision; the results were not shared with caregivers and therefore not used to alter patient treatment nor diagnosis.
- **Patient Identity:** To protect patient privacy, patient IDs were generated and assigned exclusively for this publication for patients included from the Princess Máxima Center. The translation between patient IDs and patient identity is not known outside this research group. For publicly available data, the identifiers were maintained from the metadata provided by the repository.
- **Availability of data and materials:** data will be made available via the EGA repository upon publication.
- **Code availability:** code is available upon request, and will be made publicly available through github upon publication.
- **Authors’ contributions:** CV, MPG, JdR, BT and LK conceived experiments and wrote the article. CV and MPG conceived and wrote code and performed experiments. BT and LK collected and managed patient data. MK, PW, KvB, JvdL and EH conceived experiments, wrote the article and provided clinical samples and/or information.

