## Supplementary data and figures for "Ultra-fast deep-learned pediatric CNS tumor classification during surgery"

### Contents

**Supplementary Table legends:**
Supplementary Table 1: Methylation calling accuracy.
Supplementary Table 2: Classifier training and testing over different timepoints.
Supplementary Table 3: Expected Calibration Error per class on the validation and test folds, before and after calibration.
Supplementary Table 4: Sturgeon classification scores on repeated samplings of nanopore data.
Supplementary Table 5: Simulated Nanopore sequencing runs.
Supplementary Table 6: performance on an external dataset.
Supplementary Table 7: Sturgeon performance on a pediatric cohort.

**Supplementary Figures:**
Supplementary Figure 1: F1 scores for each class at different simulated sequencing depths.
Supplementary Figure 2: F1 scores for each family at different simulated sequencing depths.
Supplementary Figure 3: Expected versus expected True Positive Rate for each different class in the validation fold prior to calibration.
Supplementary Figure 4: Expected versus expected True Positive Rate for each different class in the validation fold after temperature scaling.
Supplementary Figure 5: Expected versus expected True Positive Rate for each different class in the test fold prior to calibration.
Supplementary Figure 6: Expected versus expected True Positive Rate for each different class in the test fold after temperature scaling.
Supplementary Figure 7: True positive rate for each class when using a cutoff of 0.8.
Supplementary Figure 8: confusion matrix for pediatric samples using a cutoff of 0.95.
Supplementary Figure 9: confusion matrix for pediatric samples using a cutoff of 0.95.
Supplementary Figure 10: Robustness analysis.
Supplementary Figure 11: MinION sequencing metrics.
Supplementary Figure 12: Overlap between the nanoDx pipeline and Sturgeon classification on an external dataset.
Supplementary Figure 13: Confusion matrix for the brainstem classifier
Supplementary Figure 14: Confidence over time for the brainstem and general classifier on brainstem samples.
Supplementary Figure 15: Confidence over time for the brainstem and general classifier on non-brainstem samples.
Supplementary Figure 16: Robustness of the brainstem and general classifier for brainstem samples.
Supplementary Figure 17: Robustness of the brainstem and general classifier for non-brainstem samples.
Supplementary Figure 18: Result of four intraoperative samples using model V1.
Supplementary Figure 19: Result of four intraoperative samples using model V2.

**File: Supplementary_Table_1.xlsx**

**Supplementary Table 1: Methylation calling accuracy.** The top table shows the concordance between the EPIC profile of PMC_2 and three different methylation calling pipelines; Megalodon using a rerio CpG methylation model, Megalodon using a remora Methylation calling model and Guppy using the rerio CpG methylation calling model. n overlap shows the number of sites found that overlap with the EPIC profile. Same indicates the percentage of sites that have the same call as the binarized epic profile, false negative indicates the percentage of sites where an EPIC methylation call was called as unmethylated, False Positive indicates the percentage of site where an unmethylated EPIC call was called as methylated. The bottom table compares EPIC profiles with calls made using megalodon and the rerio model in several samples. nsites indicates the number of sites overlapping between both methods.

**File: Supplementary_Table_2.xlsx**

**Supplementary Table 2: Classifier training and testing over different timepoints.** Table includes three sheets with details regarding the classifier training on the Capper et al. dataset using different aggregations. Sheet 1 contains results when only the highest scoring subclass is taken into account, sheet 2 contains results when taking the top3 into account and sheet 3 contains results when subclasses are aggregated into the family level.Average number of CpG sites is the number of 450K array sites included for training across the simulations, true positives indicate correct outcomes, False positives indicate the number of simulations that wrongly indicated a label, False negatives indicate the number of simulations mislabeled to a class. F1-score, recall and precision are indicated for each label.

**File: Supplementary_Table_3.xlsx**

**Supplementary Table 3: Expected Calibration Error per class on the validation and test folds, before and after calibration.** Calibration was performed using temperature scaling tuned on the validation folds. This table lists the effect of calibration per class on the validation fold and on the test fold per class label.

**File: Supplementary_Table_4.xlsx**

**Supplementary Table 4: Sturgeon classification scores on repeated samplings of nanopore data.** Nanopore sequence reads were cumulatively sampled at a rate corresponding to a minION run. At each ~5 minute interval the sturgeon classifier was applied. Number of CpG sites from the 450K array and the scores per class are listed for each interval.

**File: Supplementary_Table_5.xlsx**

**Supplementary Table 5: Simulated Nanopore sequencing runs.** Nanopore sequencing runs were simulated by randomly sampling reads at a rate corresponding to the expected rate for a minION run. 100 runs were simulated per sample per timepoint, and the Sturgeon classifier was applied to each simulation. The number of correct and incorrect classifications are listed for a cutoff of 0.8 and for a cutoff of 0.95

**File: Supplementary_Table_6.xlsx**

**Supplementary Table 6: performance on an external dataset.** Sturgeon was applied to each sample, scores and outcomes were compared between the nanoDX pipeline and Sturgeon.

**File: Supplementary_Table_7.xlsx**

**Supplementary Table 7: Sturgeon performance on a pediatric cohort.** 95 EPIC profiles from a pediatric cohort, were collected. For each sample we indicate the clinical diagnosis if available. We also translated the clinical diagnosis into the closest class within the 2018 Capper et al. system if applicable. Epic arrays were submitted to the Heidelberg classifier (V11b4 unless otherwise indicated).
For each for each profile, 500 sequence experiments at each of 7 different timepoints were simulated and the Sturgeon classifier was applied. Results are listed for timepoints 1, 3 and 6. At each timepoint we indicate the number of correct outcomes at a cutoff of 0.95 (first table) or 0.8 (second table), the number of unclear outcomes, meaning the score was below the cutoff or a control class reached the threshold. Finally, we also indicate the number of wrong outcomes where an incorrect class reached the threshold. Next we indicate the most frequently found unclear or wrong class at each timepoint. In the last two tables we show results for the brainstem classifier, here samples are split based on whether their class is included in the classifier scheme.

**File: Supplementary_Table_8.xlsx**
**Supplementary Table 8:** Classes included in the brainstem classifier

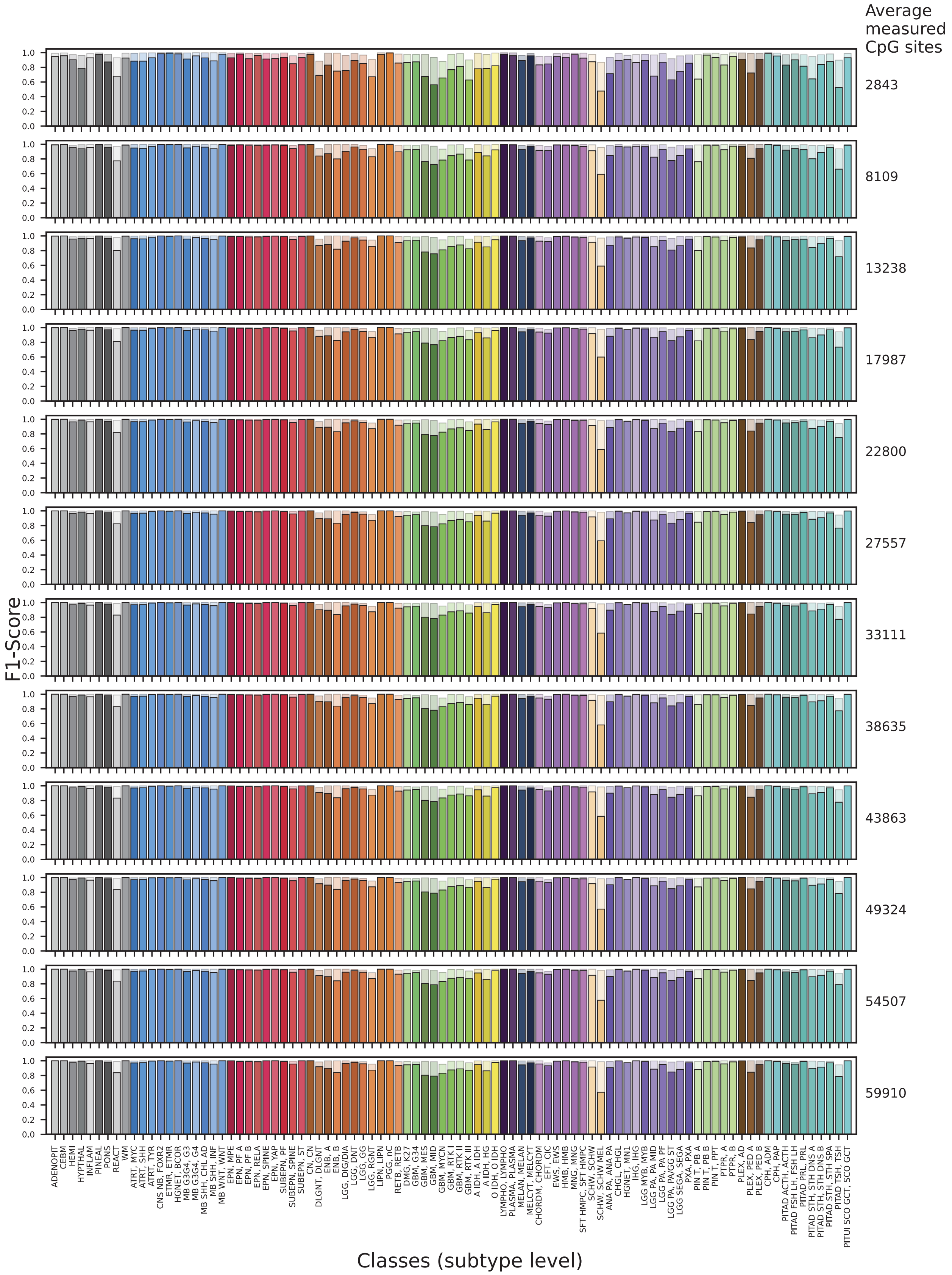


**Supplementary Figure 1: F1 scores for each class at different simulated sequencing depths.**

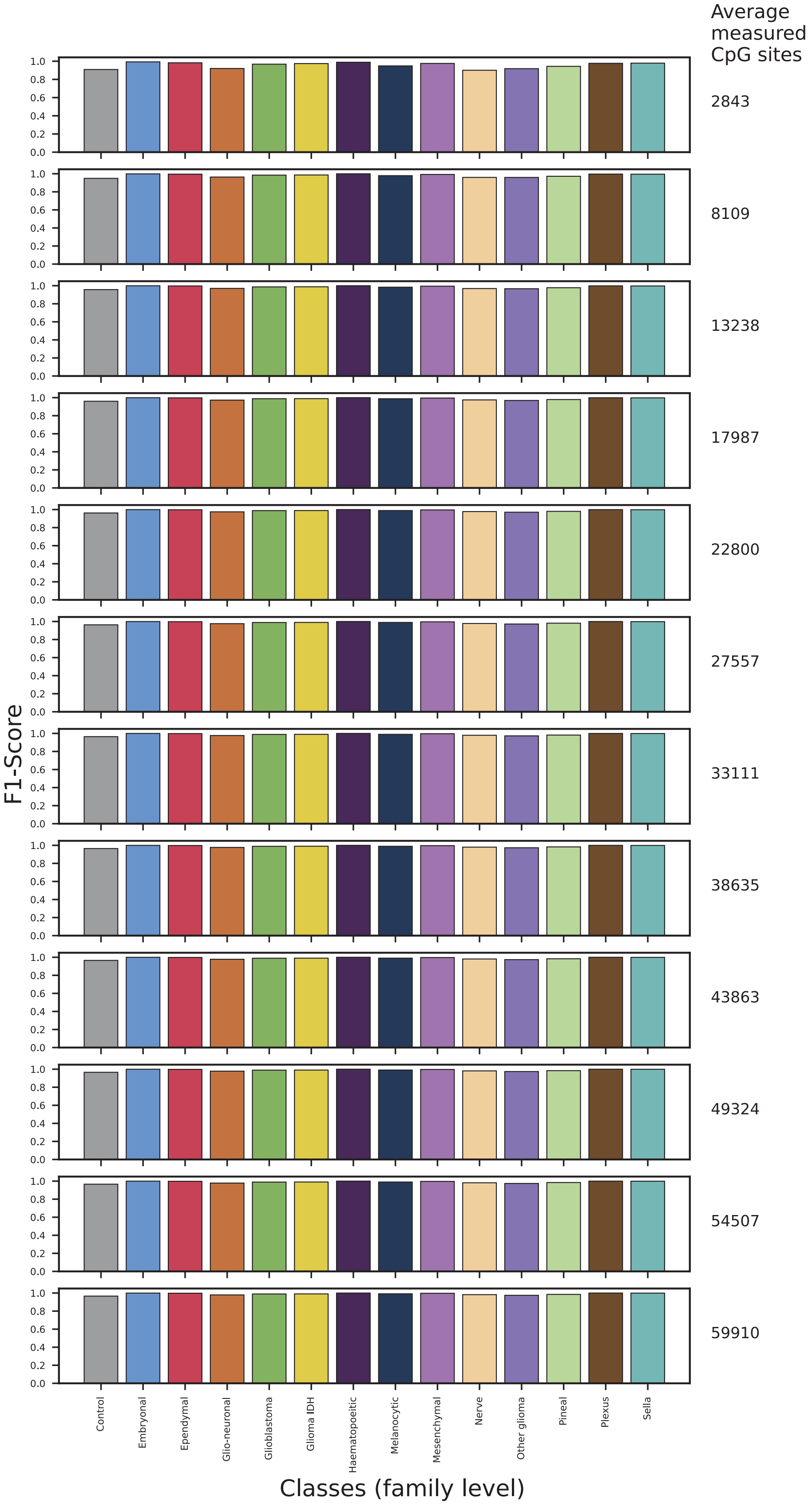


**Supplementary Figure 2: F1 scores for each family at different simulated sequencing depths.**

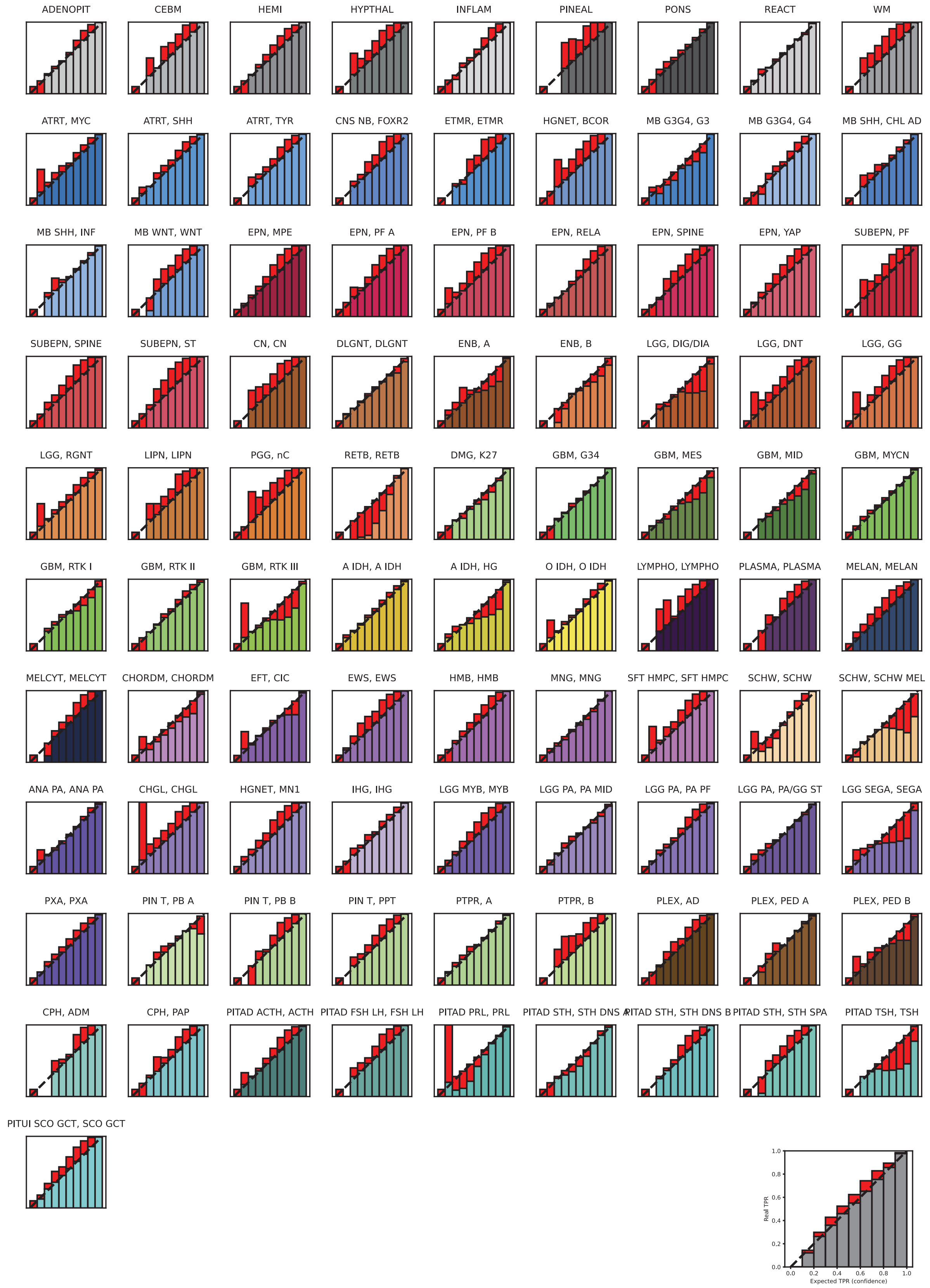


**Supplementary Figure 3: Expected versus expected True Positive Rate for each different class in the validation fold prior to calibration.**

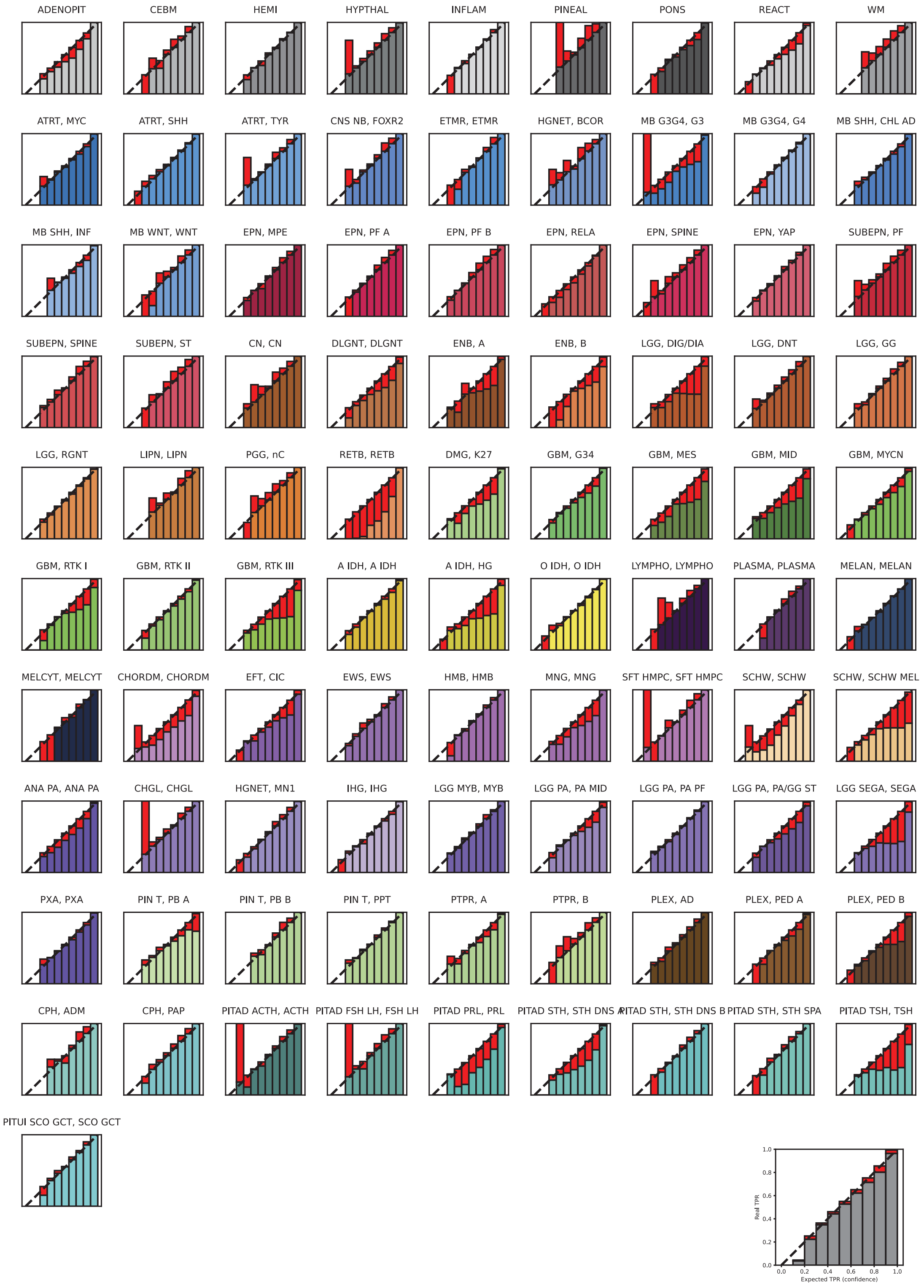


**Supplementary Figure 4: Expected versus expected True Positive Rate for each different class in the validation fold after temperature scaling.**

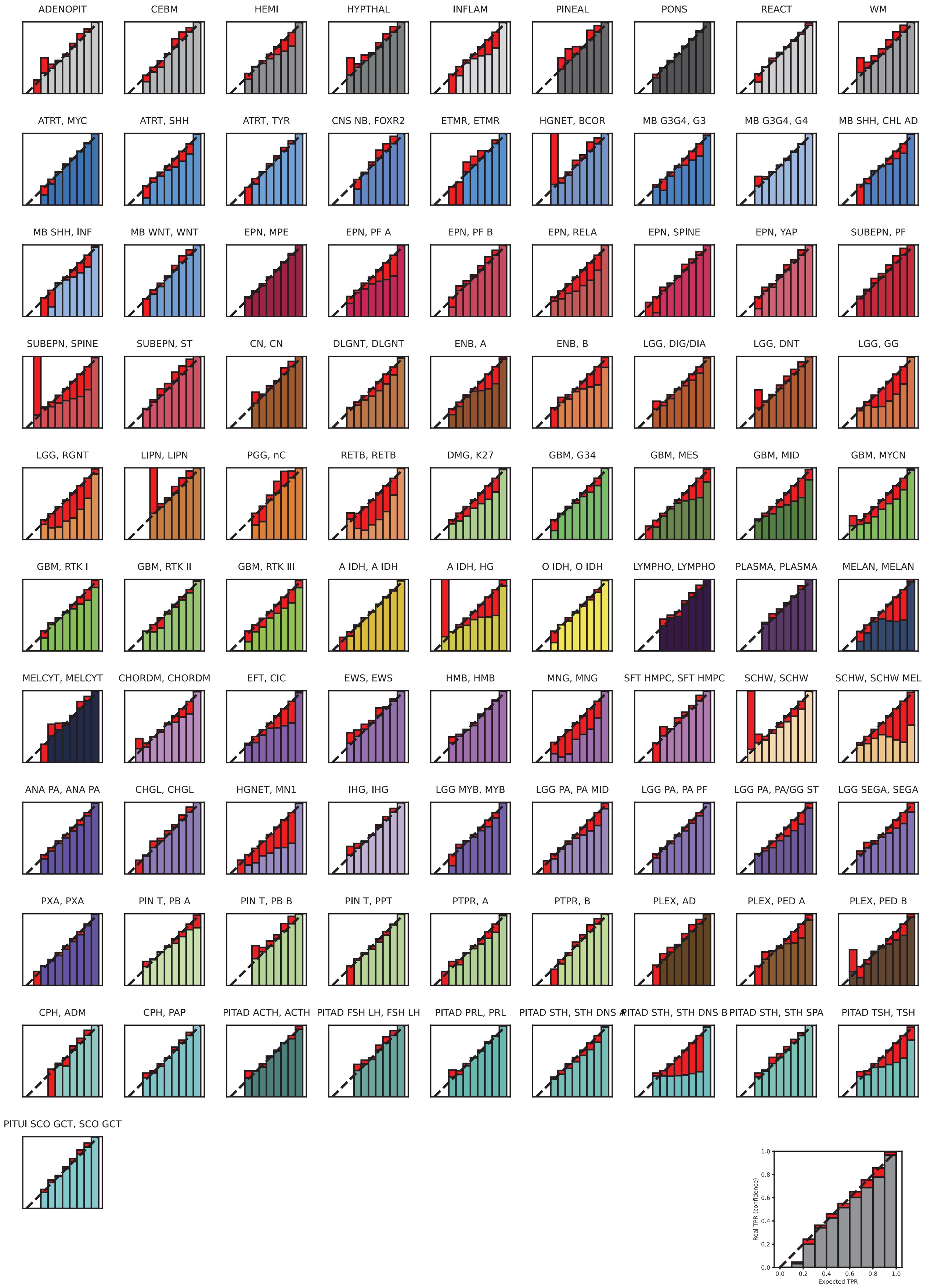


**Supplementary Figure 5: Expected versus expected True Positive Rate for each different class in the test fold prior to calibration.**

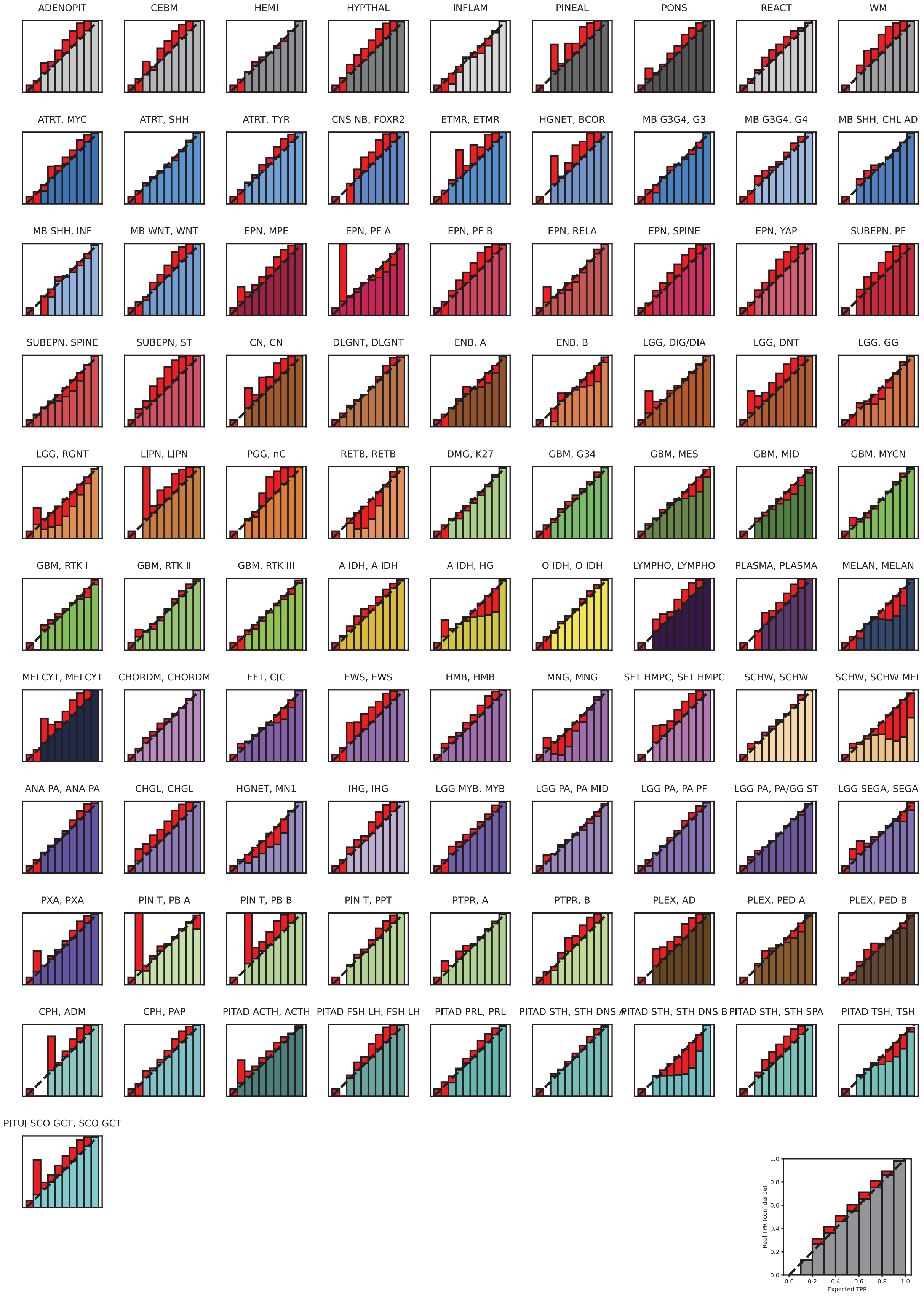


**Supplementary Figure 6: Expected versus expected True Positive Rate for each different class in the test fold after temperature scaling.**

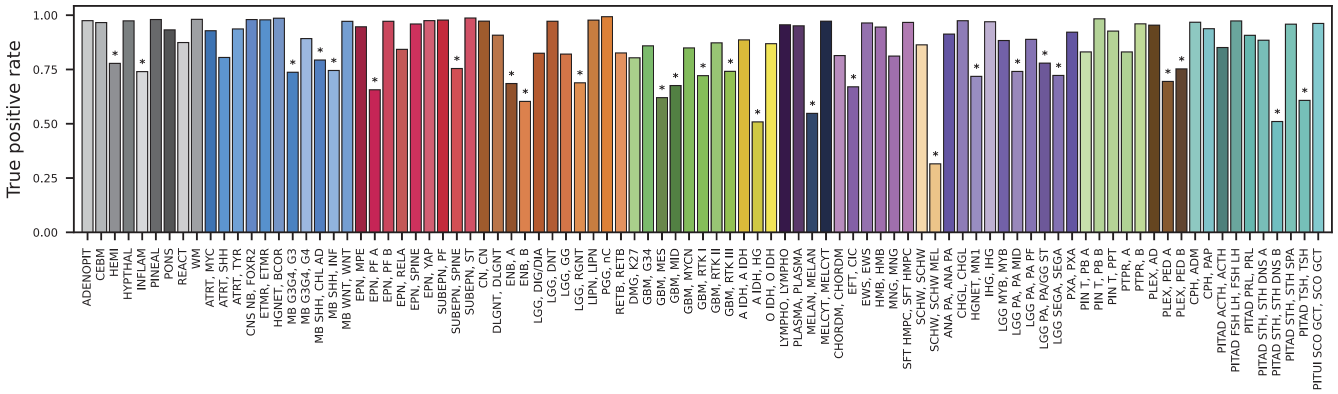


**Supplementary Figure 7: True positive rate for each class when using a cutoff of 0.8.** Asterisks indicate samples where the TPR is below 0.8.

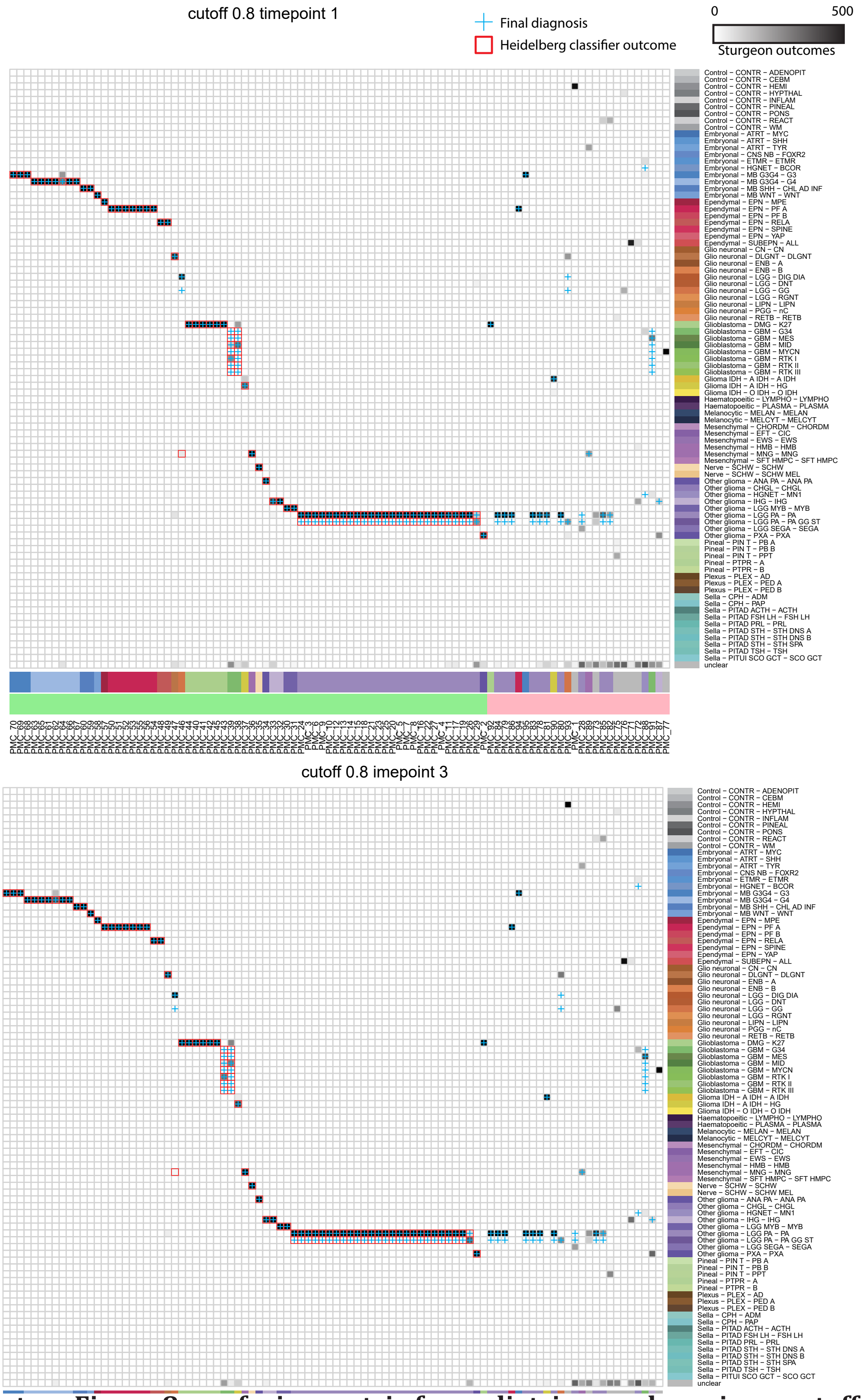


**Supplementary Figure 8: confusion matrix for pediatric samples using a cutoff of 0.95.** For each sample 500 nanopore runs were simulated at timepoint 1 and 3. The number of Sturgeon outcomes for each class is indicated in greyscale, unclear outcomes are also listed in the bottom row. Red squares indicate the Heidelberg classifier outcome (if conclusive), the blue cross indicates the clinical diagnosis.

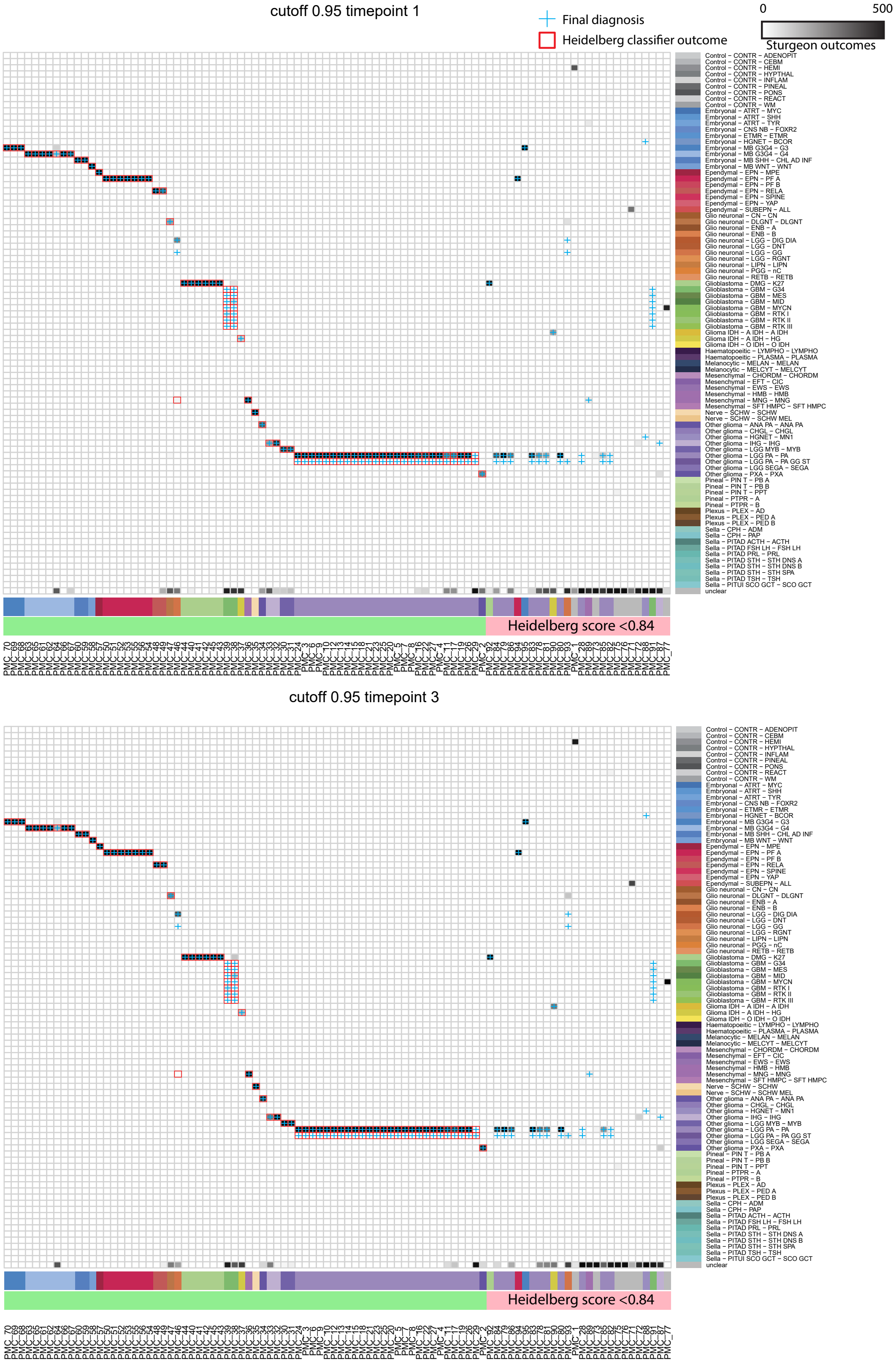


**Supplementary Figure 9: confusion matrix for pediatric samples using a cutoff of 0.95.** For each sample 500 nanopore runs were simulated at timepoint 1 and 3. The number of Sturgeon outcomes for each class is indicated in greyscale, unclear outcomes are also listed in the bottom row. Red squares indicate the Heidelberg classifier outcome (if conclusive), the blue cross indicates the clinical diagnosis.

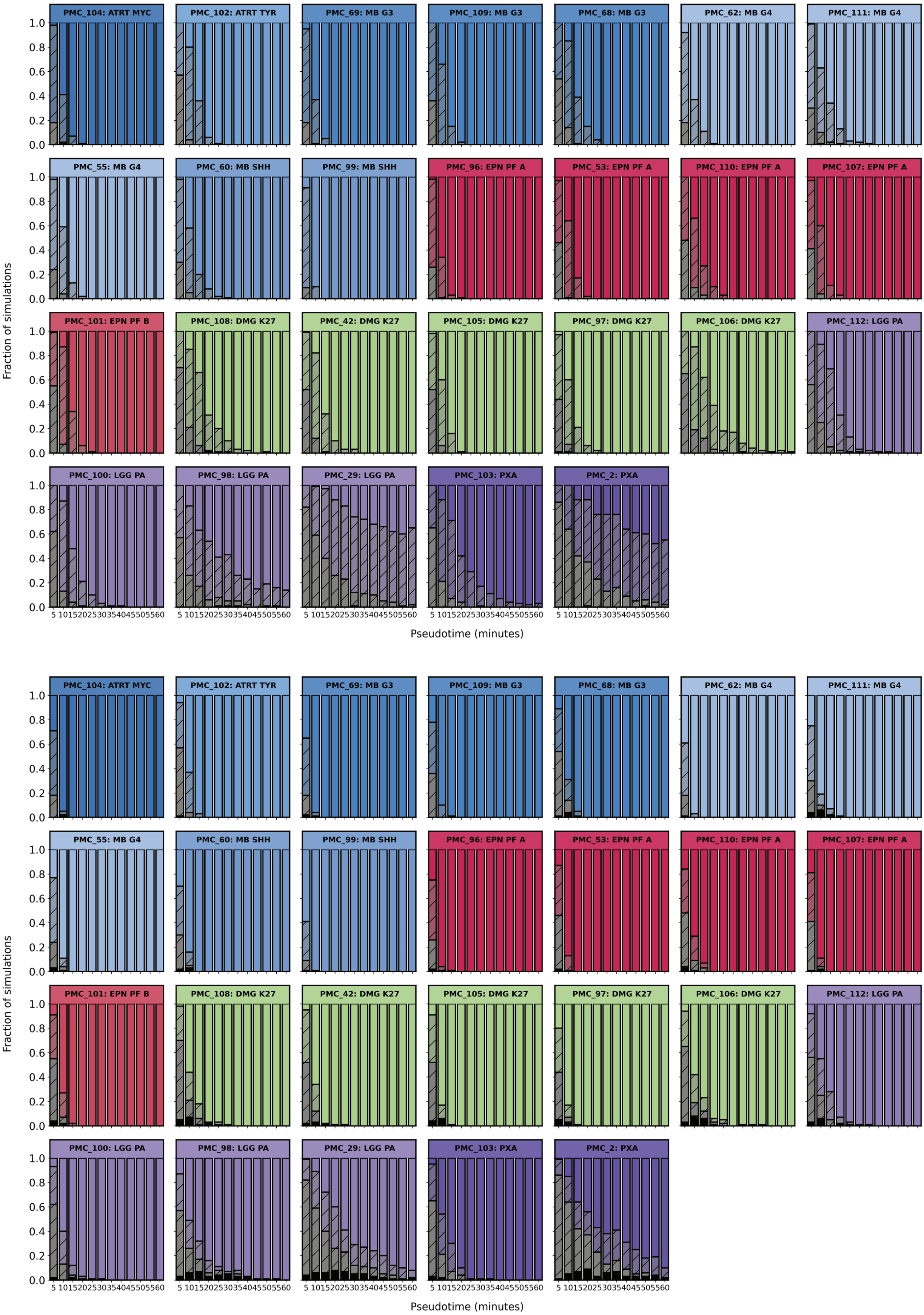


**Supplementary Figure 10: Robustness analysis.** For each sample sequence reads were randomly sampled to reflect a nanopore run at a specific duration. 100 simulations were generated for each timepoint. Colored bars indicate correct outcomes above the threshold (top 0.95, bottom 0.8), dashed colored bars indicate correct outcomes below the threshold, grey dashed bars indicate unclear outcomes and black bars indicate wrong outcomes above the treshold.

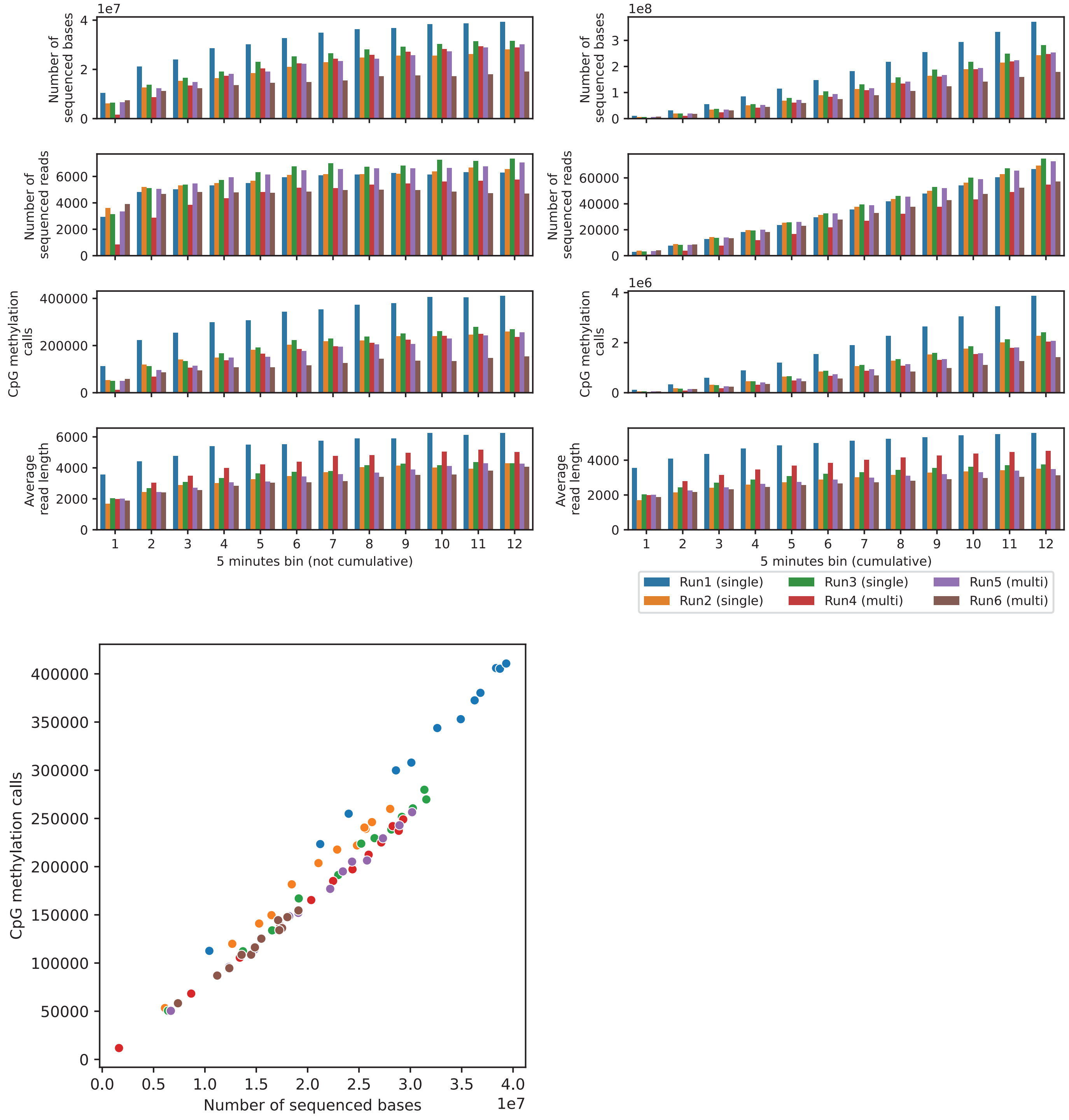


**Supplementary Figure 11: MinION sequencing metrics.** Graphs indicate the sequencing speed of minION devices used in our experiments. Indicated over time in non-cumulative (left) and cumulative (right) bins, are the number of sequenced bases, number of sequenced reads, the number of CpG methylation calls (independent of relevance to 450K arrays) and read length. Last plot indicates the number of CpG methylation calls versus the number of sequenced bases.

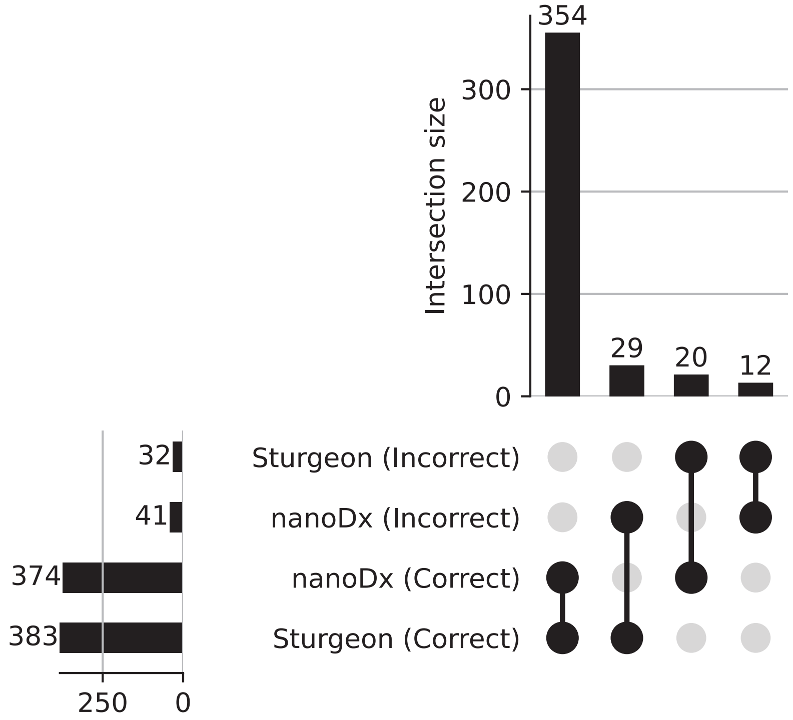


**Supplementary Figure 12: Overlap between the nanoDx pipeline and Sturgeon classification on an external dataset.**

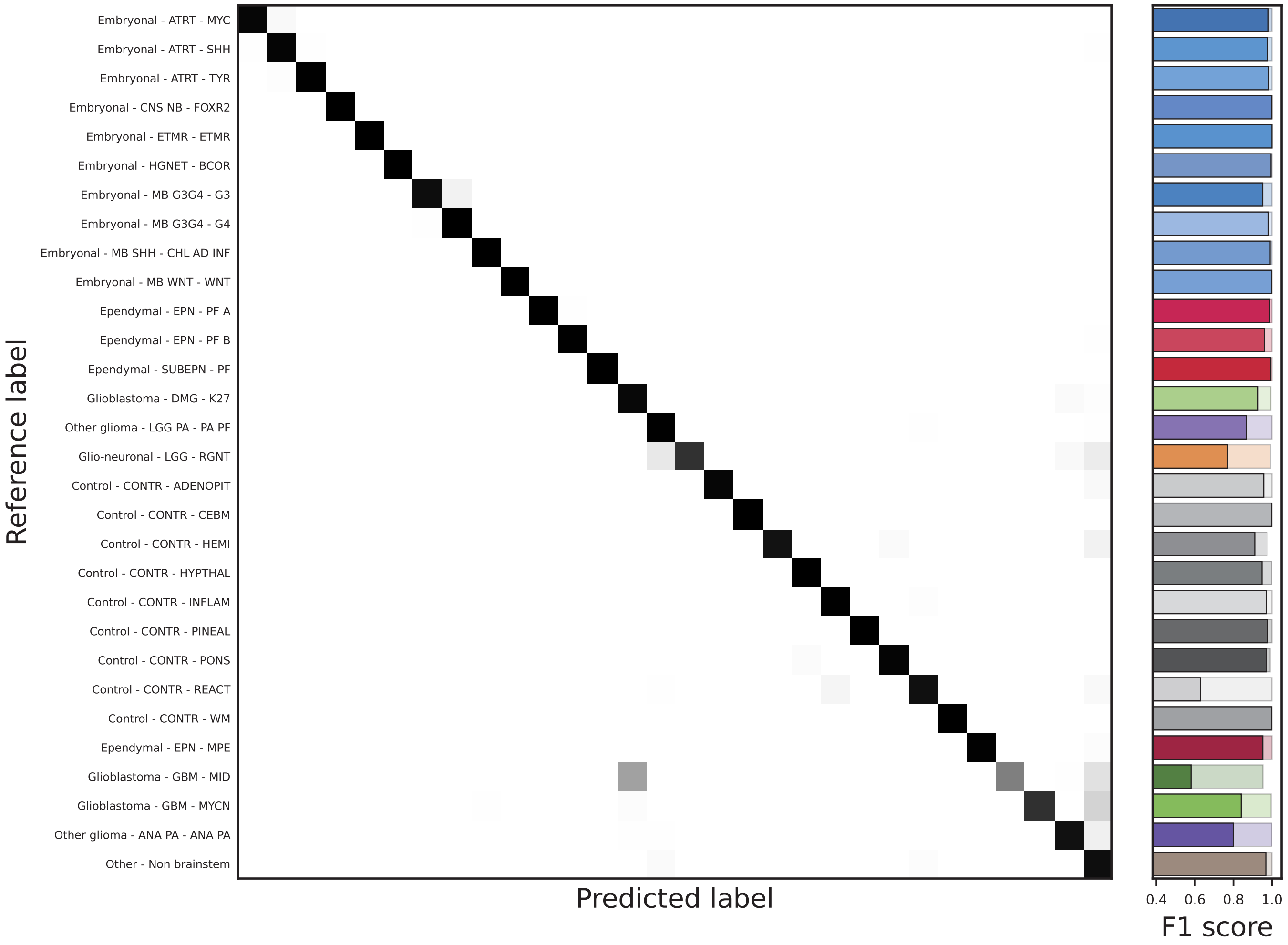


**Supplementary Figure 13: Confusion matrix for the brainstem classifier**

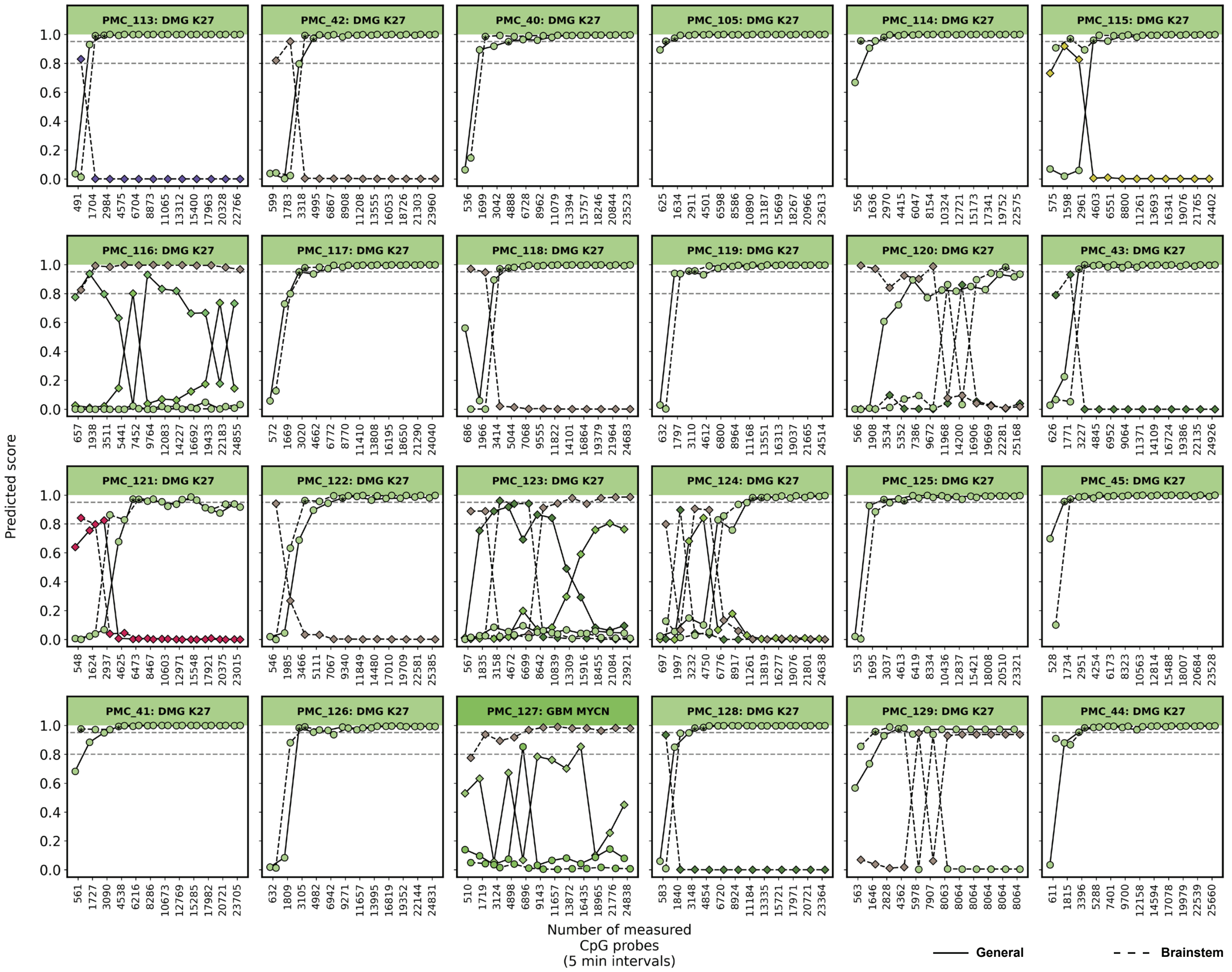


**Supplementary Figure 14: Confidence over time for the brainstem and general classifier on brainstem samples.** These plots show the confidence of the brainstem (dashed line) and general (full line) classifiers with reads accumulated at a rate expected for a minION run. Asterisks indicate the first timepoint the classification score is higher than 0.95.

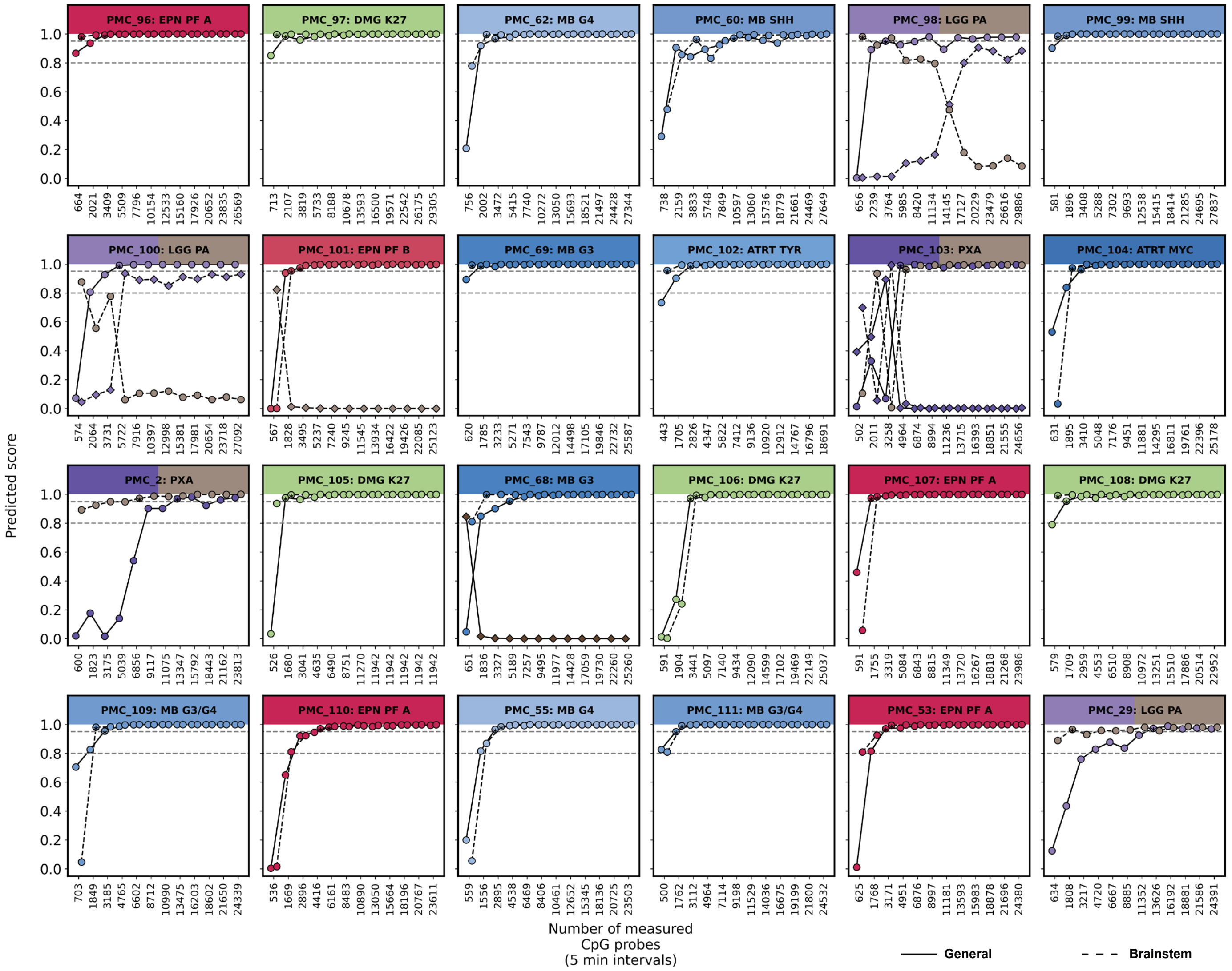


**Supplementary Figure 15: Confidence over time for the brainstem and general classifier on non-brainstem samples.** These plots show the confidence of the brainstem (dashed line) and general (full line) classifiers with reads accumulated at a rate expected for a minION run. Asterisks indicate the first timepoint the classification score is higher than 0.95.

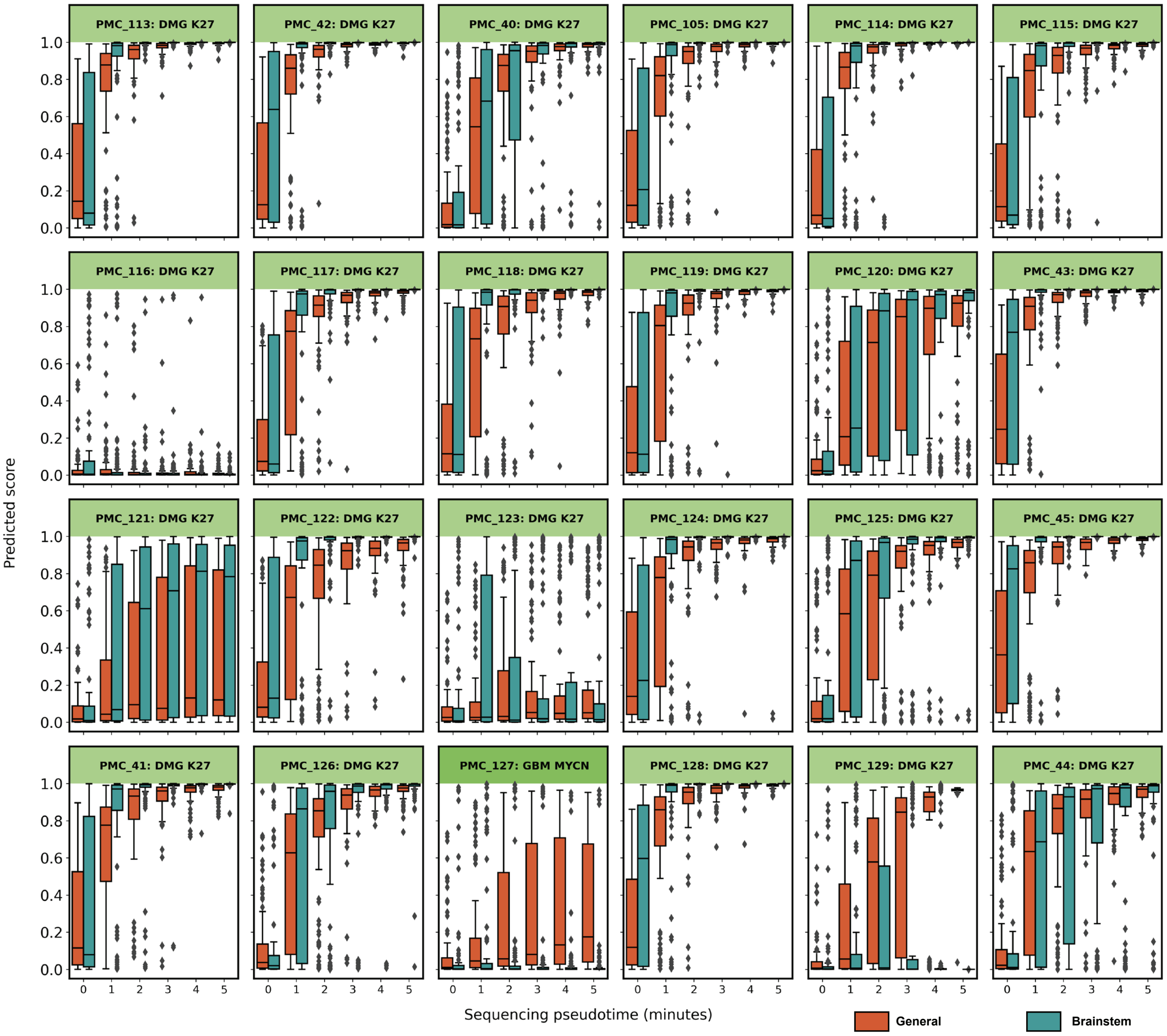


**Supplementary Figure 16: Robustness of the brainstem and general classifier for brainstem samples.** Reads were randomly sampled for each timepoint 100x and classified by the brainstem and general classifier. Boxplots indicate the score for the correct outcome in the brainstem (green) or general (orange) classifier at each timepoint.

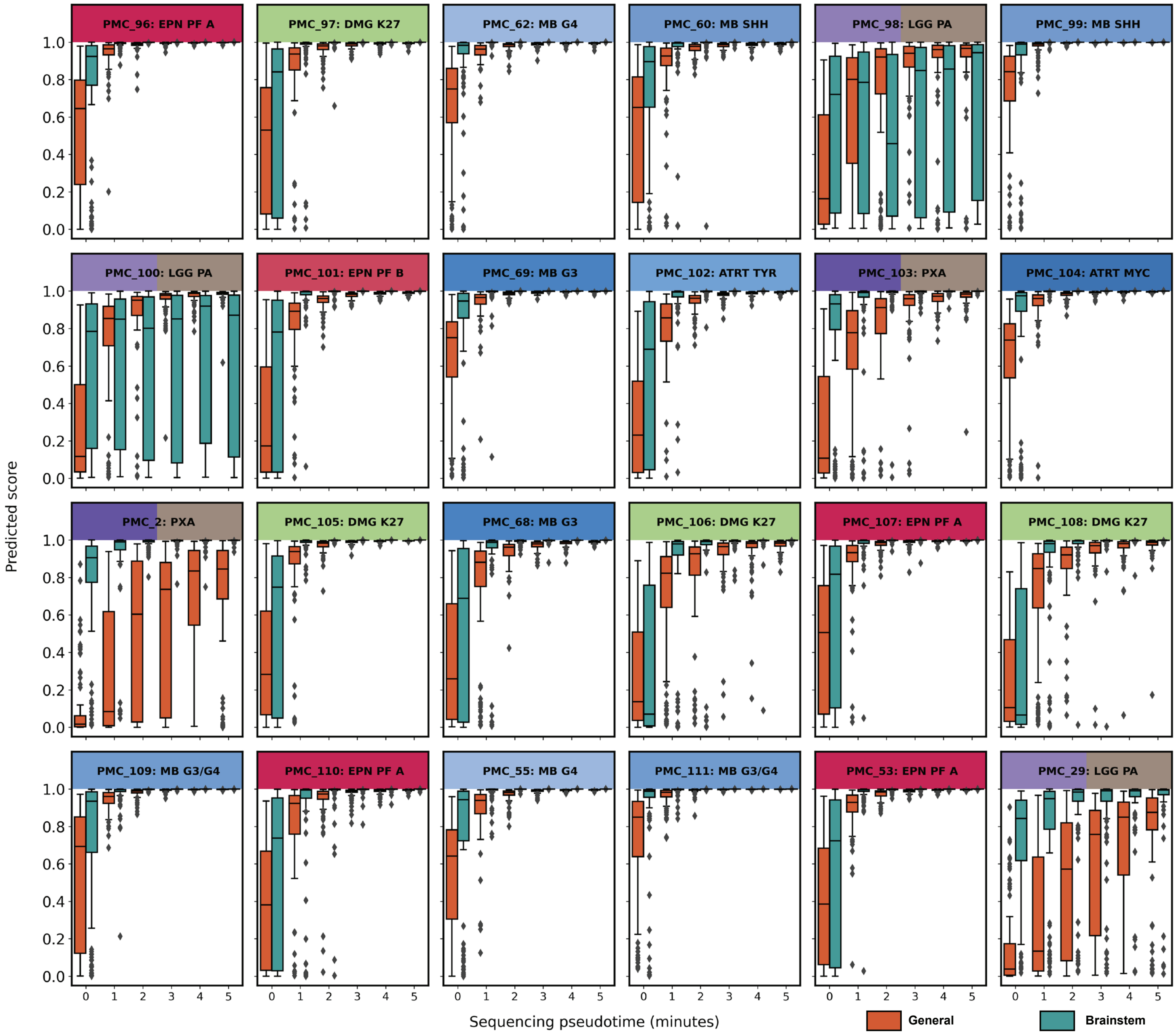


**Supplementary Figure 17: Robustness of the brainstem and general classifier for non-brainstem samples.** Reads were randomly sampled for each timepoint 100x and classified by the brainstem and general classifier. Boxplots indicate the score for the correct outcome in the brainstem (green) or general (orange) classifier at each timepoint.

PMC_130

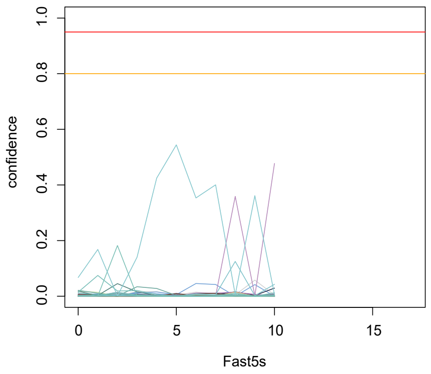


| Fast5s | time since start (minutes) | probesites | top class | confidence |
|---|---|---|---|---|
| 1 | 6,4 | 416 | Sella - PITUI SCO GCT - SCO GCT | 0,067 |
| 2 | 10,5 | 970 | Sella - PITUI SCO GCT - SCO GCT | 0,168 |
| 3 | 14,1 | 1784 | Sella - PITAD STH - STH SPA | 0,182 |
| 4 | 17,6 | 2662 | Sella - PITUI SCO GCT - SCO GCT | 0,140 |
| 5 | 20,9 | 3799 | Sella - PITUI SCO GCT - SCO GCT | 0,425 |
| 6 | 24,4 | 4891 | Sella - PITUI SCO GCT - SCO GCT | 0,544 |
| 7 | 27,6 | 6158 | Sella - PITUI SCO GCT - SCO GCT | 0,353 |
| 8 | 30,9 | 7452 | Sella - PITUI SCO GCT - SCO GCT | 0,400 |
| 9 | 34,3 | 9086 | Mesenchymal - CHORDM - CHORDM | 0,359 |
| 10 | 38,0 | 10688 | Sella - PITUI SCO GCT - SCO GCT | 0,361 |

PMC_131

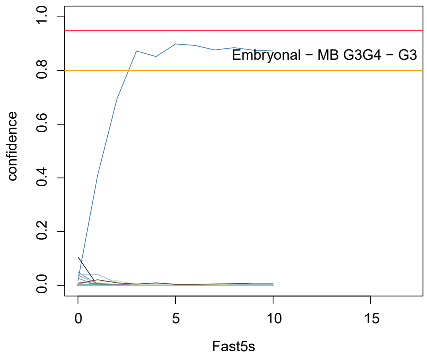


| Fast5s | time since start (minutes) | probesites | top class | confidence |
|---|---|---|---|---|
| 1 | 5,7 | 551 | Melanocytic - MELCYT - MELCYT | 0,106 |
| 2 | 9,3 | 1424 | Embryonal - MB G3G4 - G3 | 0,407 |
| 3 | 12,2 | 2464 | Embryonal - MB G3G4 - G3 | 0,695 |
| 4 | 15,1 | 3663 | Embryonal - MB G3G4 - G3 | 0,873 |
| 5 | 17,7 | 4790 | Embryonal - MB G3G4 - G3 | 0,852 |
| 6 | 20,5 | 6052 | Embryonal - MB G3G4 - G3 | 0,899 |
| 7 | 23,1 | 7543 | Embryonal - MB G3G4 - G3 | 0,894 |
| 8 | 25,8 | 9016 | Embryonal - MB G3G4 - G3 | 0,877 |
| 9 | 28,4 | 10594 | Embryonal - MB G3G4 - G3 | 0,885 |
| 10 | 31,2 | 12085 | Embryonal - MB G3G4 - G3 | 0,876 |

PMC_132

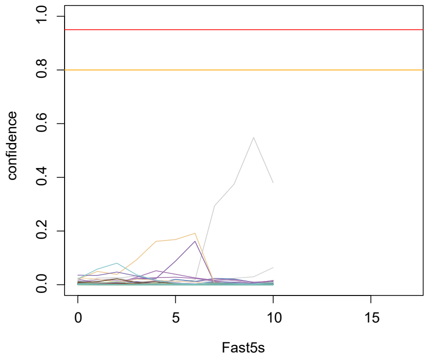


| Fast5s | time since start (minutes) | probesites | top class | confidence |
|---|---|---|---|---|
| 1 | 6,4 | 97 | Other glioma - LGG SEGA - SEGA | 0,035 |
| 2 | 10,3 | 175 | Sella - PITUI SCO GCT - SCO GCT | 0,058 |
| 3 | 13,7 | 331 | Sella - PITUI SCO GCT - SCO GCT | 0,080 |
| 4 | 16,5 | 483 | Nerve - SCHW - SCHW MEL | 0,092 |
| 5 | 19,3 | 660 | Nerve - SCHW - SCHW MEL | 0,162 |
| 6 | 21,8 | 856 | Nerve - SCHW - SCHW MEL | 0,168 |
| 7 | 24,4 | 1100 | Nerve - SCHW - SCHW MEL | 0,192 |
| 8 | 27,0 | 1352 | Control - CONTR - REACT | 0,294 |
| 9 | 29,3 | 1668 | Control - CONTR - REACT | 0,375 |
| 10 | 31,8 | 1847 | Control - CONTR - REACT | 0,549 |

PMC_133

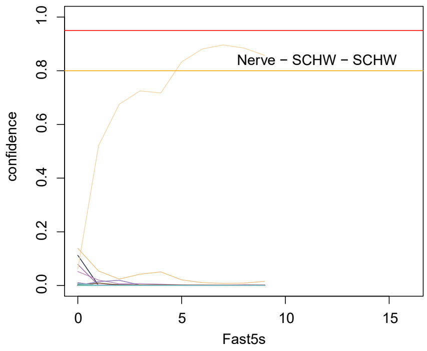


| it | time since start (minutes) | probesites | tophit | maxscore |
|---|---|---|---|---|
| 1 | 8,6 | 1035 | Nerve - SCHW - SCHW MEL | 0,139 |
| 2 | 13,9 | 2689 | Nerve - SCHW - SCHW | 0,521 |
| 3 | 18,3 | 4476 | Nerve - SCHW - SCHW | 0,676 |
| 4 | 22,5 | 6605 | Nerve - SCHW - SCHW | 0,725 |
| 5 | 26,4 | 8477 | Nerve - SCHW - SCHW | 0,717 |
| 6 | 30,2 | 10809 | Nerve - SCHW - SCHW | 0,833 |
| 7 | 34,0 | 12818 | Nerve - SCHW - SCHW | 0,881 |
| 8 | 37,7 | 15180 | Nerve - SCHW - SCHW | 0,896 |
| 9 | 41,3 | 17241 | Nerve - SCHW - SCHW | 0,885 |
| 10 | 44,9 | 19463 | Nerve - SCHW - SCHW | 0,857 |

**Supplementary Figure 18: Original results of four prospective samples.** Each sample was gathered and processed during surgery. The graph indicate the development of sturgeon scores at each Fast5 containing 4000 sequence reads. Tables indicate the time at which results were ready counted from the start of sequencing (the moment the library was loaded onto the flowcell), the number of 450K array probe sites covered at that timepoint, the highest scoring class and the score of the highest scoring class. For these experiments an older model (V1) was used.

PMC_130

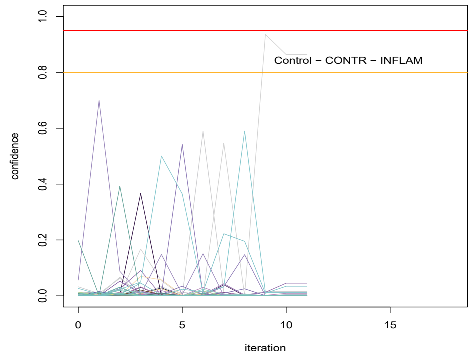


| Fast5s | Sequencing time (minutes) | probesites | highest scoring class | score |
|---|---|---|---|---|
| 1 | 6,4 | 416 | Glio-neuronal - RETB - RETB | 0,57 |
| 2 | 10,5 | 970 | Other glioma - LGG PA - PA | 0,70 |
| 3 | 14,1 | 1784 | Sella - PITAD FSH LH - FSH LH | 0,39 |
| 4 | 17,6 | 2662 | Haematopoeitic - LYMPHO - LYMI | 0,37 |
| 5 | 20,9 | 3799 | Sella - PITUI SCO GCT - SCO GCT | 0,50 |
| 6 | 24,4 | 4891 | Other glioma - CHGL - CHGL | 0,54 |
| 7 | 27,6 | 6158 | Control - CONTR - REACT | 0,59 |
| 8 | 30,9 | 7452 | Control - CONTR - INFLAM | 0,55 |
| 9 | 34,3 | 9086 | Sella - PITUI SCO GCT - SCO GCT | 0,59 |
| 10 | 38,0 | 10688 | Control - CONTR - INFLAM | 0,94 |

PMC_131

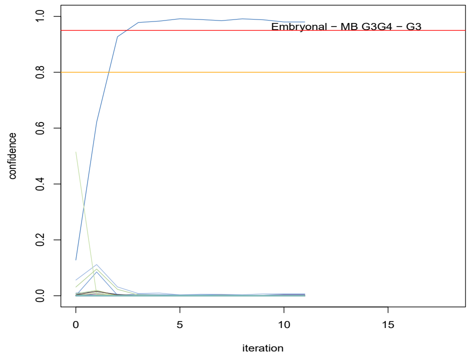


| Fast5s | Sequencing time (minutes) | probesites | highest scoring class | score |
|---|---|---|---|---|
| 1 | 5,7 | 551 | Pineal - PIN T - PB A | 0,51 |
| 2 | 9,3 | 1424 | Embryonal - MB G3G4 - G3 | 0,62 |
| 3 | 12,2 | 2464 | Embryonal - MB G3G4 - G3 | 0,93 |
| 4 | 15,1 | 3663 | Embryonal - MB G3G4 - G3 | 0,98 |
| 5 | 17,7 | 4790 | Embryonal - MB G3G4 - G3 | 0,98 |
| 6 | 20,5 | 6052 | Embryonal - MB G3G4 - G3 | 0,99 |
| 7 | 23,1 | 7543 | Embryonal - MB G3G4 - G3 | 0,99 |
| 8 | 25,8 | 9016 | Embryonal - MB G3G4 - G3 | 0,98 |
| 9 | 28,4 | 10594 | Embryonal - MB G3G4 - G3 | 0,99 |
| 10 | 31,2 | 12085 | Embryonal - MB G3G4 - G3 | 0,99 |

PMC_132

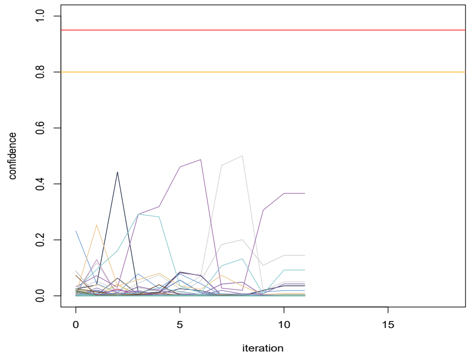


| Fast5s | Sequencing time (minutes) | probesites | highest scoring class | score |
|---|---|---|---|---|
| 1 | 6,4 | 97 | Embryonal - ATRT - TYR | 0,23 |
| 2 | 10,3 | 175 | Nerve - SCHW - SCHW MEL | 0,25 |
| 3 | 13,7 | 331 | Melanocytic - MELCYT - MELCYT | 0,44 |
| 4 | 16,5 | 483 | Sella - PITUI SCO GCT - SCO GCT | 0,29 |
| 5 | 19,3 | 660 | Mesenchymal - MNG - MNG | 0,32 |
| 6 | 21,8 | 856 | Mesenchymal - MNG - MNG | 0,46 |
| 7 | 24,4 | 1100 | Mesenchymal - MNG - MNG | 0,49 |
| 8 | 27,0 | 1352 | Control - CONTR - INFLAM | 0,47 |
| 9 | 29,3 | 1668 | Control - CONTR - INFLAM | 0,50 |
| 10 | 31,8 | 1847 | Glio-neuronal - LGG - DIG/DIA | 0,49 |

PMC_133

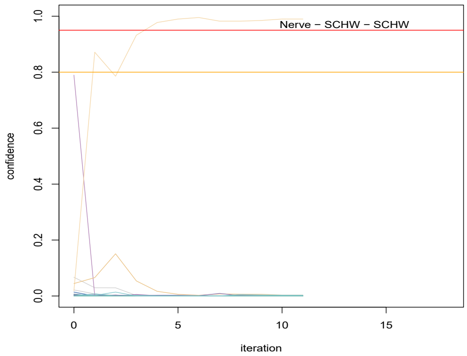


| Fast5s | Sequencing time (minutes) | probesites | highest scoring class | score |
|---|---|---|---|---|
| 1 | 8,6 | 1035 | Mesenchymal - CHORDM - CHORI | 0,79 |
| 2 | 13,9 | 2689 | Nerve - SCHW - SCHW | 0,87 |
| 3 | 18,3 | 4476 | Nerve - SCHW - SCHW | 0,79 |
| 4 | 22,5 | 6605 | Nerve - SCHW - SCHW | 0,93 |
| 5 | 26,4 | 8477 | Nerve - SCHW - SCHW | 0,98 |
| 6 | 30,2 | 10809 | Nerve - SCHW - SCHW | 0,99 |
| 7 | 34,0 | 12818 | Nerve - SCHW - SCHW | 1,00 |
| 8 | 37,7 | 15180 | Nerve - SCHW - SCHW | 0,98 |
| 9 | 41,3 | 17241 | Nerve - SCHW - SCHW | 0,98 |
| 10 | 44,9 | 19463 | Nerve - SCHW - SCHW | 0,98 |

**Supplementary Figure 19: Result of four prospective samples using the most recent model.** Each sample was gathered and processed during surgery. The graph indicate the development of sturgeon scores at each Fast5 containing 4000 sequence reads. Tables indicate the time at which results were ready counted from the start of sequencing (the moment the library was loaded onto the flowcell), the number of 450K array probe sites covered at that timepoint, the highest scoring class and the score of the highest scoring class. These are the results reproduced using the most recent model (V2).

**Supplementary video:** An entire “live sequencing run” was recorded on camera. Unfortunately the preprint server does not allow uploading this directly. It is therefore available on request by mailing
